# The global demand and potential public health impact of oral antiviral treatment stockpile for influenza pandemics

**DOI:** 10.1101/2025.02.06.25321824

**Authors:** Alvin X. Han, Katina D. Hulme, Colin A. Russell

## Abstract

Antiviral drugs are among the few countermeasures available during the critical interval between the emergence of a novel influenza pandemic and vaccine availability. Antiviral stockpiling is a key pandemic preparedness measure, yet existing stockpiling estimates vary widely and rest on outdated assumptions about healthcare-seeking behaviour and drug-specific effectiveness – limitations that the COVID-19 pandemic and recent clinical trial evidence have made untenable. We developed a multi-scale transmission model incorporating heterogeneous healthcare-seeking behaviour and the first direct clinical estimates of antiviral transmission risk reduction to estimate country-specific demand and mortality impact across four pandemic scenarios in 186 countries. We find that baloxavir marboxil (BXM), due to its transmission-reducing potential, could avert 37– 68% of mean pandemic deaths in the first epidemic wave, approximately double the impact of oseltamivir, while requiring a mean stockpile approximately 5–10% smaller (7–34% of the population, compared to 28–36% for oseltamivir). Uncertainty in viral load dynamics and transmission reduction benefits from clinical trials means that BXM’s impact could vary, but sensitivity analyses consistently suggest that BXM is likely to be more effective than oseltamivir. Under limited drug availability, priority should be given to treatment over post-exposure prophylaxis. Although drug rationing for high-mortality populations (e.g. elderly) can substantially reduce BXM demand, doing so leads to greater total pandemic deaths. Critically, each week of delay in initiating antiviral distribution erodes impact by up to 3% of averted deaths, meaning that antiviral stockpiles must be accompanied by rapid deployment infrastructure to deliver their potential impact.

**Significance Statement:** Pandemic preparedness requires strategic stockpiling of antiviral drugs but existing stockpile estimates are based on oversimplified assumptions about healthcare-seeking behavior and drug effectiveness. This study develops a new mathematical model incorporating realistic healthcare-seeking patterns and drug-specific transmission risk reduction when considering antiviral stockpile planning for influenza pandemics across 186 countries. We find that baloxavir marboxil could prevent nearly twice as many deaths as oseltamivir while requiring smaller stockpiles. Under limited drug supply, treatment should be prioritised over prophylaxis, and age-based rationing for high-mortality populations would not reduce total pandemic deaths. These insights provide evidence-based guidance for national antiviral drugs stockpiling strategies as part of preparedness efforts for future influenza pandemics.

## Introduction

Influenza A viruses (IAV) remain an emerging threat to human health owing to persistent spillover risk of novel IAV subtypes from reservoir species. Given their antiviral activity against diverse IAV subtypes and the convenience of administration, orally or inhalation administered influenza antiviral drugs are potentially effective countermeasures to mitigate the impact of a nascent influenza pandemic prior to the availability of a targeted vaccine. Several approved drugs could be considered for pandemic preparedness stockpiling, including adamantanes, M2 ion-channel blockers preventing the initiation of viral replication, neuraminidase inhibitors such as oseltamivir, laninamivir and zanamivir that attenuates the release of progeny virions from infected cells, polymerase inhibitors such as baloxavir marboxil (BXM) and favipiravir that blocks viral replication, as well as umifenovir, a fusion inhibitor that prevents virus fusion to host cells. Although laninamivir and zanamivir are currently recommended for post-exposure prophylactic use, they, alongside favirpiravir and umifenovir, are not recommended by the World Health Organization for treatment of both severe and non-severe influenza infections given their limited effects in reducing illness duration (1–4), and thus unlikely to form the primary antiviral stockpile. Adamantanes are also unlikely to be useful as the primary antiviral drug due to the high propensity of developing resistance among adamantane-treated individuals (5). Although the emergence and spread of oseltamivir and BXM resistance are also of concern, prompt treatment with either drug reduces symptom duration and risk of severe disease outcomes (5, 6). Both treatments are also effective post-exposure prophylactic drugs that lower infection risk (2). Importantly, treatment with BXM has recently been demonstrated to provide a significant clinical benefit in preventing onward transmissions (7).

Mathematical modelling is a useful tool to rationally design evidence-based antiviral stockpiling options amidst uncertainties about the future influenza pandemic. However, existing modelling evidence is inconsistent and even conflicting owing to differences in modelling objectives, approaches and assumptions, making it challenging to meaningfully synthesise for public health planning. For example, focusing on treatment demand estimation, a neuraminidase inhibitor stockpile covering ~30% of the population was estimated to be sufficient to treat all symptomatic individuals infected by the pandemic virus that seek care in the US during a nascent pandemic (8). Conversely, a separate study estimated a much larger antiviral demand covering 40-144% of the Canadian population to account for additional contingencies such as treating non-specific infections that also present influenza-like illness (9). Several studies separately used simpler decision-tree approaches to design stockpile size options aimed at maximizing economic benefits. In Singapore, the cost-effective treatment-only oseltamivir stockpile size was estimated at 40-60% per-capita (10) while Siddiqui et al. suggest that stockpiling oseltamivir for the entire population of the UK for treatment is more prudent since the attack rate of the future pandemic virus is unknown (11). In contrast, a dynamic transmission model applied to ten countries, including Singapore and UK, estimated a much smaller treatment-only oseltamivir stockpile size of 15-25% per-capita to minimise mortality rates and economic costs while also finding little benefit in giving antiviral prophylaxis to exposed contacts (12). Other studies found that restricting antivirals for treatment is ineffective at limiting the impact of the pandemic virus (13), or that spread of the pandemic virus can only be constrained under widespread distribution of post-exposure prophylaxis (PEP), covering 20-50% of exposed contacts (14) or that a nascent pandemic can even be contained at its source if >80% of exposed contacts were given prophylaxis (15, 16).

Previous studies would likely now reach different conclusions in light of the COVID-19 pandemic and recent clinical data. First, all of the aforementioned studies assumed that at least 70% of symptomatic infections could be treated within two days of symptom onset. While symptomatic individuals might readily seek care during a pandemic (17), healthcare-seeking behaviour, including the time to seeking treatment since symptom onset, is heterogeneous, even during a pandemic (18), depending on sociodemographic factors and disease severity (19). Similarly, studies proposing mass distribution of PEP have assumed that countries can swiftly mobilise extensive and near-complete contact tracing but this level of capacity during the COVID-19 pandemic was rare even in high-resource countries (20, 21). Several studies also assume that the infectiousness of infected individuals treated with oseltamivir is reduced by >60% (12, 15, 16, 22), a figure derived from post-hoc analyses (23) of clinical trial data that assessed oseltamivir as PEP for preventing infection in uninfected household contacts, and not transmission risk reduction by treating infected index patients (24, 25). This assumption is likely a substantial overestimate: BXM reduces infectious viral load more effectively than oseltamivir (6), yet direct measurement in a randomised clinical trial (CENTERSTONE) shows BXM reduces onward transmission from treated index patients by only 29% (7) – implying that the transmission reducing effect of oseltamivir is lower still. Despite their differences in scope and approach, these studies share a reliance on simplified, largely pre-pandemic assumptions about healthcare-seeking behaviour and antiviral effectiveness that have since been challenged by empirical evidence.

In this study, we developed a mathematical modelling framework to estimate the potential antiviral demand (defined here as the number of treatment courses that could be utilised under different access assumptions) and associated impact on burden reduction when using either BXM or oseltamivir to mitigate the initial wave of an influenza pandemic (see Methods and SI Appendix). Our deterministic model simulates how within-host viral dynamics would change as a result of antiviral administration and its consequent impact on between-host transmission dynamics. We used the latest estimates of transmission risk reduction of antivirals based on direct measurements in clinical trials, and accounted for the impact of varying delays in treatment administration since infection. We applied our model to 186 countries where we have reliable demography and age-stratified contact pattern estimates, providing country-specific demand and impact estimates under the same modelling framework. We did not incorporate the distribution of vaccines in our model. While emerging technologies may expedite vaccine development, their effectiveness, the timing and manner of vaccine rollout in each country during a nascent pandemic are uncertain. Since vaccines strongly influence impact and demand of antivirals, we opted to exclude this variable. That said, we had previously showed that if vaccines are available, expanding vaccination coverage will be the more efficient option over mass test-and-treatment with antivirals to lowering disease burden (26). We also estimated the additional antiviral demand and potential benefits in distributing PEP to household contacts. Finally, we quantified how the spread of antiviral resistance could potentially impact antiviral demand and burden reduction. Under the reference scenario, we assumed that 95% of all symptomatic individuals would ideally seek test-and-treat within two days after symptom onset, with no restrictions to availability and access to antivirals. We also assumed that country-wide distribution of antivirals could ideally begin one week after pandemic initiation in the country. We opted not to account for non-pharmaceutical interventions (NPIs) in our model. While NPIs can lower antiviral demand and augment disease burden reduction, it is challenging to assume an appropriate effectiveness of NPIs, to which modelling results will be highly sensitive, as they depend on the type of NPIs, time-dependent variation, as well as influences owing to political, governance, cultural, behavioural and socioeconomic context (27, 28).

## Results

### Potential antiviral treatment demand and mean deaths averted

Under the idealised reference scenario, we find that BXM broadly doubles the percentage of mean pandemic deaths averted relative to oseltamivir (Figure 1) with fewer mean treatment courses (Figure 2) across all pandemic scenarios and countries (Figure 3). The median percentage of mean pandemic deaths averted across all countries depends on the severity of the pandemic scenario. BXM could avert 68% of mean deaths in the relatively mild 2009 A/H1N1-like pandemic (interquartile range (IQR) = 62-100%; 13-34 deaths averted per million people) but only 48% in a 1968 A/H3N2-like pandemic (IQR = 45-52%; 194-329 mean deaths averted per million people), 37% in a COVID-19-like pandemic (IQR = 35-40%; 843-2,588 mean deaths averted per million people) and 44% in a 1918 A/H1N1-like pandemic (IQR = 42-45%; 2,798-2,993 mean deaths averted per million people). The corresponding median mean per-capita demand of BXM is 7% for a 2009 A/H1N1-like pandemic (IQR = 5-11%; 410-902 million courses of BXM as per the estimated 8.2 billion world population in 2024(29)), 24% for a 1968 A/H3N2-like pandemic (IQR = 21-25%; 1.7-2.0 billion courses), 34% for a COVID-19-like pandemic (IQR = 33-36%; 2.7-3.0 billion courses) and 28% for a 1918 A/H1N1-like pandemic (IQR = 25-29%; 2.0-2.4 billion courses). In contrast, oseltamivir averts median mean deaths by 35% (IQR = 31-41%), 27% (IQR = 26-28%), 23% (IQR = 23-24%) and 25% (IQR = 25-26%) for the 2009 A/H1N1-like, 1968 A/H3N2-like, COVID-19-like and 1918 A/H1N1-like influenza pandemics respectively. Yet, the median mean oseltamivir demand is ~5-10% greater than BXM, correspondingly amounting to 32% (IQR = 28-33%; 2.3-2.7 billion courses of oseltamivir), 28% (IQR = 25-30%; 2.0-2.5 billion courses), 36% (IQR = 35-37%; 2.9-3.0 billion courses) and 32% (IQR = 28-33%; 2.3-2.7 billion courses) per-capita for the four pandemic scenarios.

**Figure 1:**
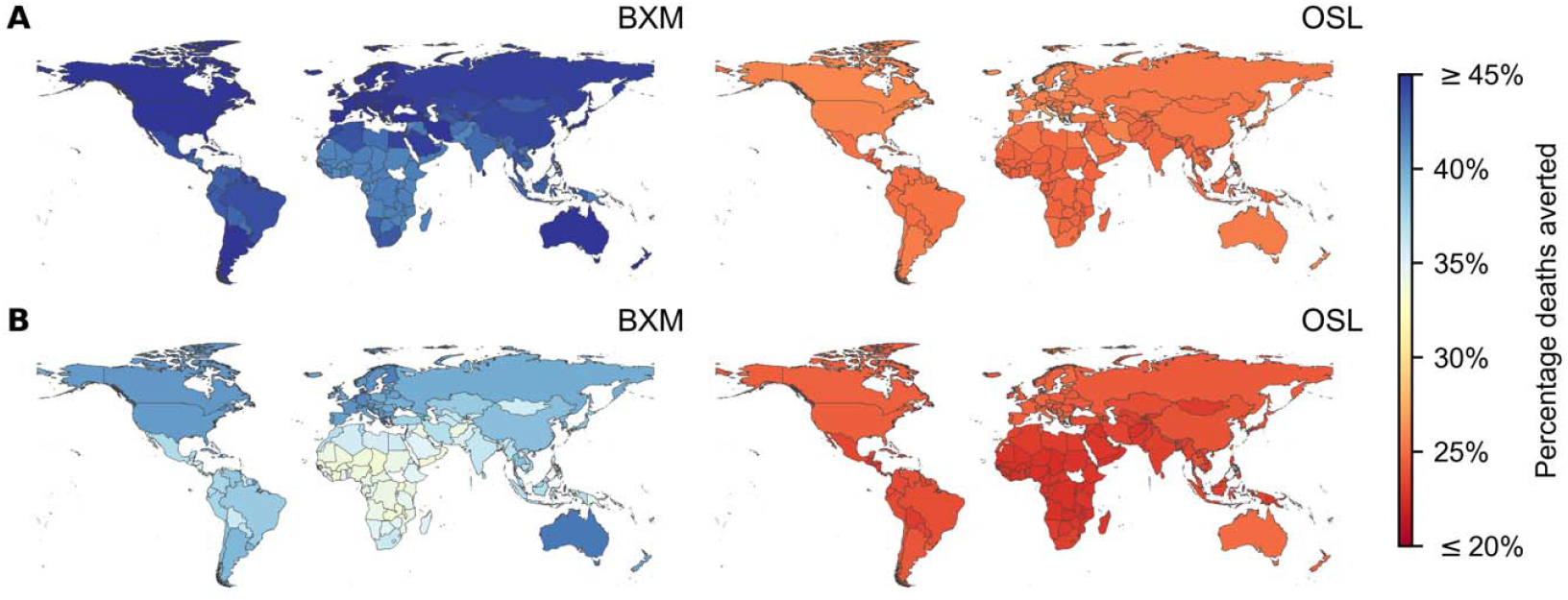
Mean percentage deaths averted by test-and-treat with oral antivirals in 186 countries under idealised assumptions. Baloxavir marboxil (BXM; left column) or oseltamivir (OSL; right column) was administered to all individuals that were positively diagnosed, using a rapid test with 70% test sensitivity, within two days of symptom onset. All symptomatic individuals were assumed to have sought testing within one day after symptom onset on average. Distribution of oral antivirals for treatment began one week after the pandemic was initiated in the country with ten infections. (**A**) 1918 A/H1N1 pandemic (=2.0). (**B**) Hypothetical influenza pandemic with COVID-19-like disease burden (=3.0) but generation interval akin to seasonal influenza viruses.

**Figure 2:**
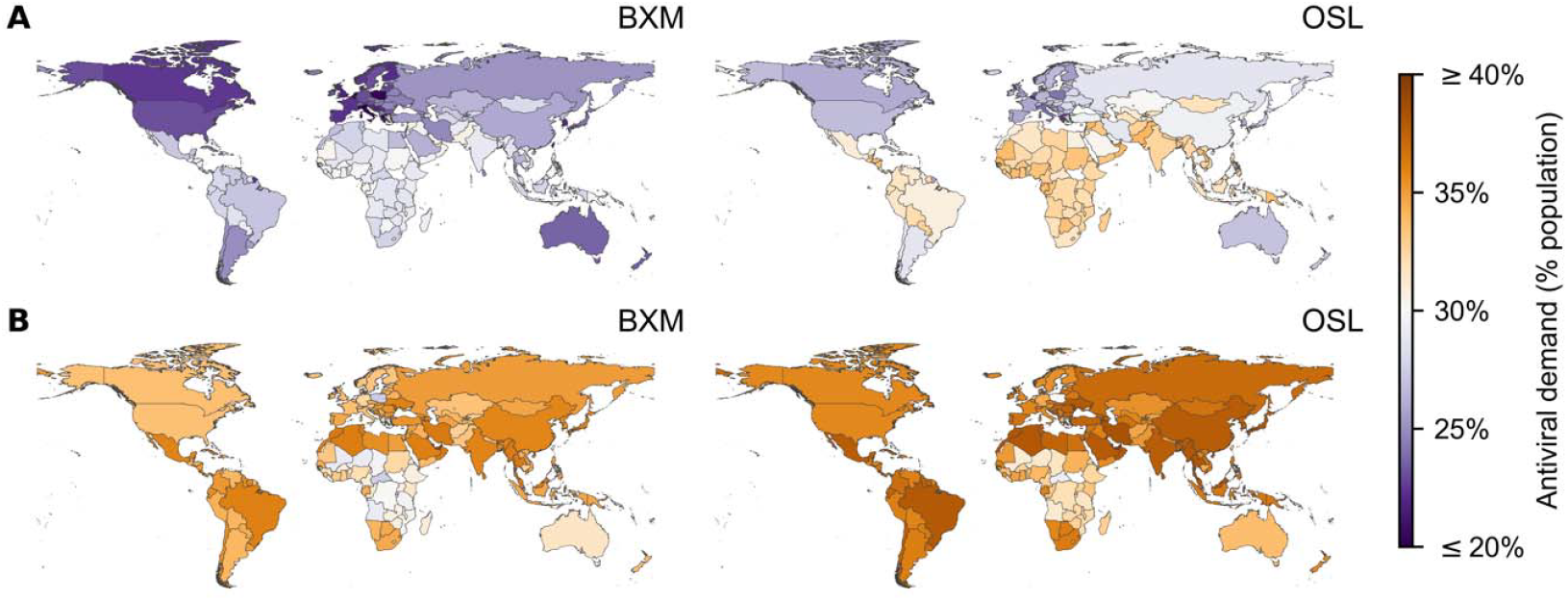
Potential treatment oral antiviral demand of 186 countries under idealised assumptions. Baloxavir marboxil (BXM; left column) or oseltamivir (OSL; right column) was administered to all individuals that were positively diagnosed, using a rapid test with 70% test sensitivity, within two days of symptom onset. All symptomatic individuals were assumed to have sought testing within one day after symptom onset on average. Distribution of oral antivirals for treatment began one week after the pandemic was initiated in the country with ten infections. (**A**) 1918 A/H1N1 pandemic (=2.0). (**B**) Hypothetical influenza pandemic with COVID-19-like disease burden (=3.0) but generation interval akin to seasonal influenza viruses.

**Figure 3:**
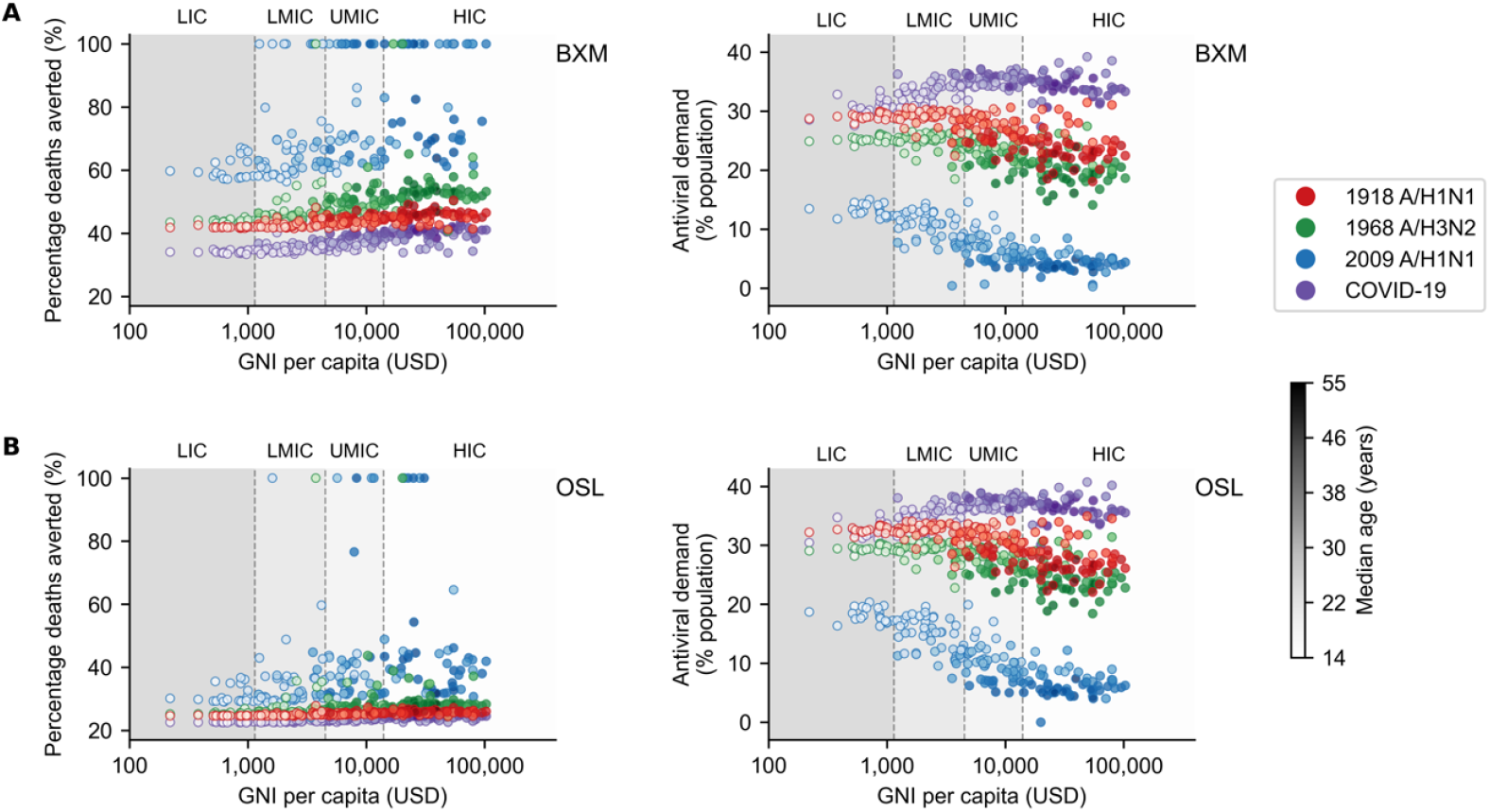
Demographic and economic covariates to treatment oral antiviral demand and impact. Mean percentage deaths averted (left column) and potential treatment oral antiviral demand (right column) for each country (circle, shading denotes median age of country’s population) under different pandemic scenarios (denoted by colour of circles; red, 1918 A/H1N1; green, 1968 A/H3N2; blue, 2009 A/H1N1; and purple, COVID-19-like) are plotted against their gross national income (GNI) per capita is US dollars (USD) in 2023. Background shading denotes the range of GNI per capita distinguishing between different country income groups as classified by the World Bank in 2024 (low income country, LIC; lower-middle income country, LMIC; upper-middle income country, UMIC; and high income country, HIC). Antiviral drugs were administered to all individuals that were positively diagnosed, using a rapid test with 70% test sensitivity, within two days of symptom onset. All symptomatic individuals were assumed to have sought testing within one day after symptom onset on average. Distribution of oral antivirals for treatment began one week after the pandemic was initiated in the country with ten infections. (**A**) Baloxavir marboxil. (**B**) Oseltamivir.

At the country level, owing to their generally younger demography, the potential per-capita mean impact on disease burden by either antiviral is lower in lower-middle and low income countries while requiring up to four times more treatment courses relative to high income countries (Figure 3), except for the COVID-19-like pandemic scenario which, unlike historical influenza pandemics, impacted younger individuals to a lesser extent owing to their lower susceptibility to SARS-CoV-2 infection, and in turn, reducing the estimated mean antiviral treatment demand in lower income countries. Table S3 tabulates the estimated potential mean antiviral treatment demand, mean rapid testing demand and the corresponding mean deaths averted for each individual country across all four simulated pandemic scenarios.

We also performed sensitivity analyses to account for uncertainty surrounding key parameters, including: (1) parameters simulating within-host viral dynamics, (2) transmission risk reduction by BXM where the 95% confidence interval was estimated at 12-45% (7), and (3) the probability of asymptomatic infection, which we varied between 0.05 and 0.40. First, our deterministic modelling framework estimated the mean impact and demand of antiviral distribution and in doing so, we had generated maximum likelihood estimates of within-host parameters based on clinical trial data (6). However, there was considerable variability in changes to viral titers as a result of BXM or OSL administration in the clinical trial. To account for this variability, we performed Bayesian Markov Chain Monte Carlo sampling to obtain a posterior distribution of within-host parameters, fitted jointly to the viral load data observed in placebo, BXM and OSL arms in the clinical trial (see SI Appendix). We then propagated the posterior draws (n=500) to the between-host transmission model, simulating for the distribution of BXM and OSL in the United States under the reference pandemic scenarios. BXM showed wider 95% credible intervals in mean percentage of deaths averted for the relatively less severe pandemics (A/H3N2-like and 2009 A/H1N1-like) compared to the more severe scenarios (1918 A/H1N1-like and COVID-19-like) (Table S4). While credible intervals overlapped between BXM and OSL in all instances, BXM exhibited non-trivial impact on deaths averted (i.e. more than zero) for all simulated pandemic scenarios and skewed toward higher mean percentages deaths averted, suggesting greater impact in spite of the uncertainty.

Second, with greater transmission risk reduction by BXM, mean percentage deaths averted expectedly tend to increase with decreasing antiviral demand (Figure S1). However, the marginal changes to mean percentage deaths averted and antiviral demand depends on the pandemic simulated, with more diffuse changes estimated for milder pandemics. That said, even if BXM only conferred 12% transmission risk reduction, under the reference scenario, BXM could avert 46-100% (IQR) of mean deaths (13-33 mean deaths averted per million people) for 2009 A/H1N1-like pandemic, 38% in a 1968 A/H3N2-like pandemic (IQR = 36-39%; 161-253 mean deaths averted per million people), 34% in a COVID-19-like pandemic (IQR = 33-35%; 788-2,287 mean deaths averted per million people), and 36% in a 1918 A/H1N1-like pandemic (IQR = 35-37%; 2,301-2,463 mean deaths averted per million people). Finally, both mean percentage deaths averted and antiviral demand also expectedly decrease with higher probability of asymptomatic infection (Figure S2 and S3). BXM tends to result in larger decreases in mean percentage deaths averted than oseltamivir since fewer symptomatic infections are tested for treatment with higher asymptomatic infection rates.

### Impact of treatment delay, distribution strategies, post-exposure prophylaxis and antiviral resistance

We subsequently investigated how pandemic death reduction and the corresponding demand for BXM, the potentially more impactful antiviral drug, would change by deviations in reality from the idealised reference scenario. We used a Bayesian multilevel model to estimate the joint impact of these deviations (Figure 4). First, distribution of test and treatment across the country is often delayed beyond one week since pandemic initiation in the country. Sufficient epidemiological evidence must accumulate before a pandemic emergency can be ascertained. Additionally, resource, logistical and public health infrastructure constraints, especially in low-resourced countries, can further impede timely test-and-treat interventions. Excluding pandemic- and country-specific effects, percentage of mean deaths averted by BXM decreases by 0.9% (95% highest posterior density (HPD) = 0.1-1.6%) for every week of delay to test-and-treatment distribution since the first infections in the country. This decrease is further exacerbated under pandemic scenarios with faster growth rates, for instance each week of distribution delay under a COVID-19-like pandemic scenario decreases percentage of mean deaths averted by 2.9% (95% HPD = 2.8-3.0%). Without prompt delivery and accessibility to test-and-treatment, the theoretical utility of a pandemic oral antiviral stockpile is greatly diminished (26).

**Figure 4:**
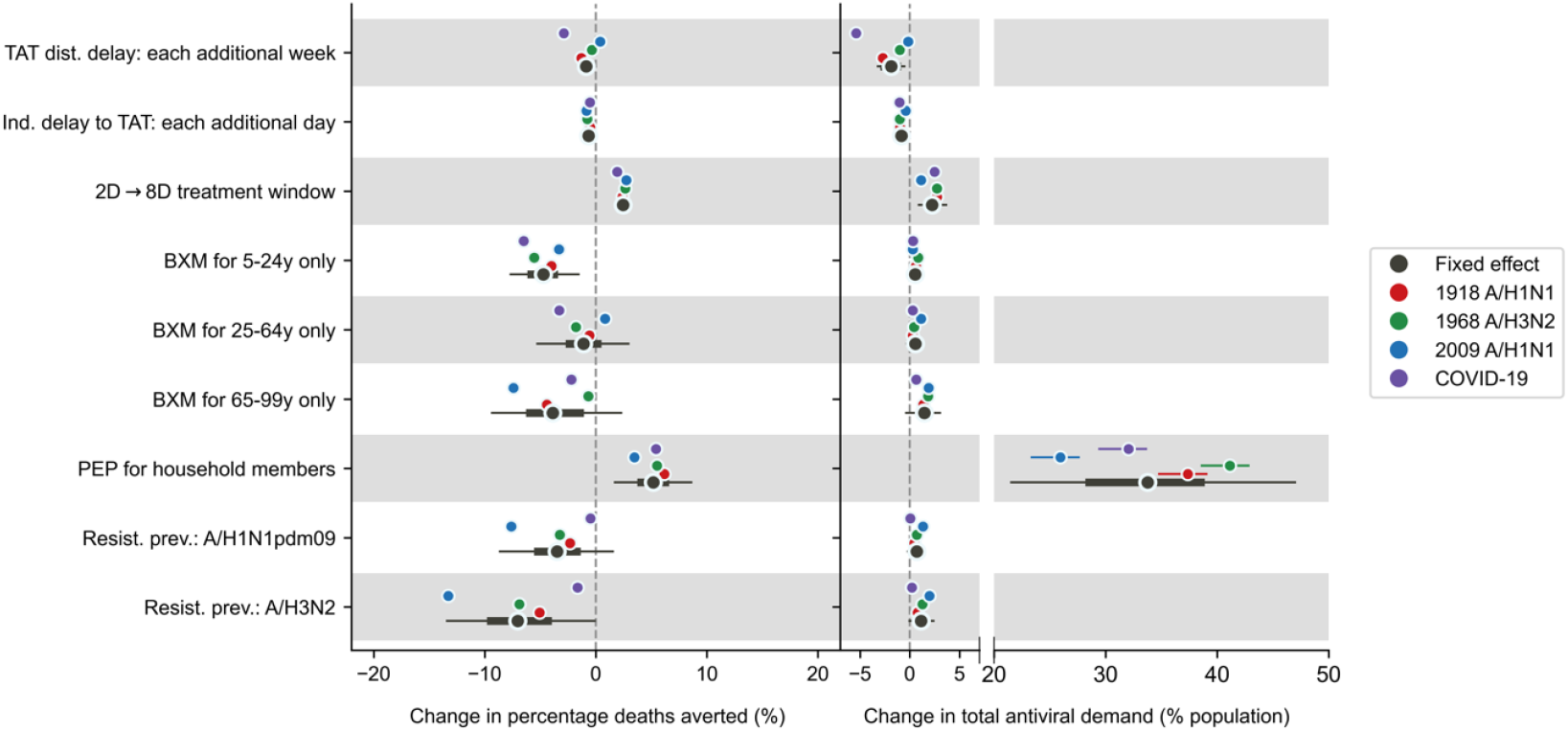
Joint effects of treatment delays, distribution strategies, post-exposure prophylaxis and antiviral resistance on mean percentage deaths averted and total antiviral demand. Relative to the idealised reference scenario (i.e. antivirals are swiftly distributed to patients across the country one week after the first infection in the country, with no limits on access and availability of tests and antivirals; >95% of all symptomatic individuals are treated within two days after symptom onset if positively diagnosed by a rapid diagnostic test; No emergence and spread of BXM-resistant viruses), mean posterior changes in mean percentage deaths averted (left plot) and total antiviral (i.e. BXM and oseltamivir) demand (right plot) across pandemics and countries are plotted as black circles (i.e. fixed effects) while the thick and thin lines denote the corresponding 67% and 95% highest posterior density intervals respectively. The corresponding pandemic scenario-adjusted posterior effects are plotted alongside (posterior mean and 95% highest posterior density interval denoted by colour of circle and line respectively; red, 1918 A/H1N1; green, 1968 A/H3N2; blue, 2009 A/H1N1; and purple, COVID-19-like).

It is also unlikely that most infected persons would seek test-and-treatment within two days of symptom onset on average given heterogenous healthcare-seeking behaviour across the population (18, 19). For each additional day of delay in mean time to treatment since symptom onset across the population, the population-level reduction in mean deaths is expected to decrease by 0.7% (95% HPD = 0.5-0.8%). To mitigate the diminishing impact of individual treatment delay, we considered extending the current indicated BXM treatment use of within two days post-symptom onset to eight days. Leveraging on BXM’s rapid inhibition of viral replication (6), additionally treating individuals who sought treatment after two days post-symptom onset could theoretically lower their infectiousness (Figure S4) such that population-level percentage mean deaths averted would increase by 2.4% (95% HPD = 1.8-3.0%) while requiring 2.3% (95% HPD = 0.9-3.6%) more per-capita courses of BXM.

Age-based rationing is a common resource saving strategy under limited drug availability. We estimated how overall pandemic deaths and BXM demand would change, relative to the reference scenario of no age restriction, if BXM was differently restricted for individuals aged 5– 24 years, 25–64 years, and those ≥ 65 years of age only while other age groups were allotted oseltamivir instead. Rationing BXM for different age groups leads only to trivial changes in total antiviral (i.e. BXM and oseltamivir) demand (i.e. age group, 95% HPD: 5–24 years, −0.1-1.2%; 25–64 years, −0.3-1.4%; ≥ 65 years, −0.3-3.0%). However, mean BXM demand could approximately be halved if rationed for individuals aged 5–24 years, lowered by ~60% if given to 25–64 years only and further reduced by ~96% if only seniors ≥ 65 years are given BXM, with variation depending on the pandemic and the country (Table S5).

Changes to percentage mean pandemic deaths averted depends on the age group allotted BXM (Figure 4). If BXM is given to younger individuals aged 5–24 years only, percentage mean pandemic deaths averted decreases by 4.7% (95% HPD = 1.6-7.7%). If BXM is restricted to individuals aged 25–64 years, changes to percentage mean pandemic deaths averted strongly depends on pandemic-specific effects with trivial country-level effects despite the variation in age demography between countries (Figure S5). For example, if BXM is distributed to 25–64 years only, percentage mean deaths averted would decrease by 3.3% (95% HPD = 3.0-3.6%) under COVID-19-like pandemic scenario but increase by 0.8% (95% HPD = 0.5-1.2%) during the 2009 A/H1N1-like pandemic which disproportionately affected middle-aged adults (30) (Figure 4). If BXM is allotted to older individuals ≥ 65 years of age only, decrease in percentage mean deaths averted depends on the pandemic scenario (i.e. 1918 A/H1N1-like, 95% HPD = −4.9-4.0%; 1968 A/H3N2-like, 95% HPD = −1.1-0.2%; 2009 A/H1N1-like, 95% HPD = −7.9-7.0%; COVID-19-like, 95% HPD = −2.7-1.8%; Figure 4).

Although BXM is effective as PEP (31), swift and extensive contact tracing for PEP distribution can be challenging to implement during a pandemic (20, 21). Under the ideal assumption of same-day BXM administration to household contacts upon the identification of the index patient, excluding pandemic- or country-level effects, providing BXM as PEP on top of treatment increases mean pandemic deaths averted by 5.2% (95% HPD = 1.7-8.6%) while incurring additional per-capita BXM demand by 33.8% (95% HPD = 21.6-47.0%). Changes in BXM impact and demand due to PEP varies by country (Figure S5–7) where lower-income countries, which tend to have larger mean household sizes, would benefit from PEP distribution when mean household size is ≥ 4 members, further increasing mean pandemic deaths averted by up to ~6%. However, realizing this benefit would require up to ~50% more per-capita mean BXM demand.

Multiple studies reported that seasonal IAVs readily acquire treatment-emergent BXM-resistant mutations without a loss in viral fitness (6, 32–36), especially among children, albeit at different rates of emergence (32, 33, 36), which have been shown to be transmissible between ferrets (36, 37) and potentially between humans (34, 36). We assessed the degree to which widespread transmission of resistance mutations could potentially diminish the impact and demand of BXM, assuming treatment-emergent resistance prevalence at average levels observed for seasonal A/H1N1pdm09 and A/H3N2 viruses. Under the higher prevalence of treatment-emergent resistance akin to treatment of A/H3N2 infections, excluding pandemic- and country-level effects, mean pandemic deaths averted by BXM could potentially decrease by 7.0% (95% HPD = 0.1-13.4%) relative to no resistance emergence. Pandemics with slower growth rates would more likely lead to wider resistance spread, as evidenced by lower total percentage of mean pandemic deaths averted (Figure 4). Conversely, resistance spread under A/H1N1pdm09 resistance prevalence yields trivial fixed effects on mean pandemic deaths averted (95% HPD = −8.6-1.5%) and is instead strongly dependent on the pandemic scenario (Figure 4) with generally trivial country-level effects (Figure S5) despite the age dependency of resistance emergence rates. Assuming that rapid testing does not distinguish between drug-sensitive and −resistant viruses for treatment administration, changes in mean BXM treatment demand are trivial under both treatment-emergent resistance prevalence levels (i.e. A/H3N2, 95% HPD = 0.0-2.0%; A/H1N1pdm09, 95% HPD = −0.2-1.5%).

## Discussion

To investigate how heterogeneous time to test and treatment with antivirals since infection impact influenza virus infectiousness and transmission dynamics, we developed a novel multi-scale and multi-type model that links heterogeneous within-host infectiousness owing to variation in timing of antiviral administration to between-host influenza transmission dynamics. This is also the first study that incorporates recent clinical estimates of transmission risk reduction by antiviral treatment (7). We estimated the potential country-specific influenza antiviral demand for either BXM or oseltamivir and their corresponding impact in reducing pandemic deaths during a nascent influenza pandemic for 186 countries under the same age-structured transmission model, resolving difficulties in either synthesizing or extrapolating findings from previous modeling studies, which have focused on either one or a handful of countries based on used outdated assumptions, to inform potential influenza antiviral stockpiling options that fit country-specific needs.

Across antiviral drugs, an estimated 20-40% of the population of each country could meaningfully benefit from treatment during a novel influenza pandemic, with lower-income countries generally experiencing greater need. At these levels of drug use, treatment-only antiviral strategies could avert >20% of expected pandemic deaths during the first epidemic wave. Because of its demonstrated benefits in reducing transmission, BXM is both more effective and more economical in treatment courses: its distribution doubles pandemic death reduction relative to oseltamivir, with the potential to avert 37-68% of mean pandemic deaths, while requiring ~5-10% fewer courses of drugs. However, due to existing patent and cost constraints on BXM, oseltamivir remains the more accessible option for lower income countries.

The accessibility constraints of BXM are also complicated by the lack of transparency of national antiviral stockpiling strategies and the compounding challenges posed by opaque drug pricing. Unless global health actors, in particular high-income countries, muster the political will to address the many challenges underlying global health inequity (38, 39), cooperation between regional states could present opportunities to increase affordable access to BXM, alongside other pandemic medicines, pre-pandemic vaccines, and emerging vaccine development technologies, before the next influenza pandemic through pooled procurement mechanisms and/or technology transfer agreements (40). Nonetheless, procuring enough antivirals to sufficiently meet demand does not necessarily confer the aforementioned impact estimates unless similar investments are made to public health infrastructure to ensure prompt drug distribution across a country upon pandemic initiation, with enough testing resources to identify most symptomatic individuals shortly after infection.

The urgency of pre-pandemic stockpiling is underscored by the severe time-sensitivity of antiviral effectiveness. Our modelling estimates that each week of delay in initiating country-wide test-and-treat distribution reduces mean deaths averted by approximately 1% under moderate pandemic conditions, rising to nearly 3% per week under a COVID-19-like pandemic scenario where rapid epidemic growth leaves little margin for logistical delays. A stockpile that exists on paper but cannot be rapidly deployed therefore provides a false sense of preparedness. Procuring antivirals is necessary but insufficient; equivalent investment in the public health infrastructure required for prompt detection, testing, and distribution is inseparable from the stockpile’s value.

Under limited availability, age-based rationing of BXM while providing oseltamivir to the rest of the population can substantially lower BXM demand even though total antiviral courses needed will likely remain unchanged. Rationing for individuals with greater risk of mortality (i.e. the elderly) or those with greater mobility, and thus more likely to spread the virus (i.e. young individuals), would both lead to more pandemic deaths, the magnitude of which depends on the pandemic scenario. However, rationing BXM for the elderly could lead to far greater reduction in BXM demand by >90%. Cost-effectiveness analyses should be performed to determine the appropriateness of age-based rationing under different pandemic scenarios and country-specific drug prices but are beyond the scope of this work.

Besides treatment, providing antivirals as PEP to exposed contacts could confer additional benefits in reducing disease burden. Household members of index patients are potentially easier contact tracing targets for PEP distribution and a recent modelling study found that combining treatment of the index patient and PEP for exposed household members could yield considerable reduction in burden within individual households (41). However, given the heightened risk of infection during a pandemic across the population, independent of country-level effects, distributing BXM to household contacts would only lead to modest population-level increase in mean pandemic deaths averted by ~5% while increasing mean BXM demand by >30%. PEP would only additionally reduce disease burden in settings where the mean household size is ≥4. Under limited drug availability and/or below the mean household size of four, prioritizing antiviral drugs for treatment as opposed to PEP is more effective and treatment course-efficient considering population-level outcomes.

Finally, it is important to consider the degree to which antiviral resistance attenuates impact as it may be more prudent to limit BXM to individuals that would most likely derive clinical benefit from treatment as opposed to mass antiviral distribution. When assuming mean treatment-emergent prevalence of BXM resistant mutations was similar to seasonal A/H3N2 infections, percentage reduction in mean pandemic deaths under the spread of BXM resistance was still substantial, estimated to be >24% compared to no intervention. As such, in spite of resistance spread, mass distributing BXM for treatment could still lead to substantial life-saving benefits. Various strategies have been proposed to potentially suppress the spread of resistance, such as cycling the distribution of two different antiviral drugs (42) or combining them for treatment (43, 44). Previous modelling work showed that either strategy could work well to suppress resistance spread if the probability of resistance emergence of the primary antiviral drug is <5% (42), which could be the case if treatment-emergent resistance prevalence of the pandemic virus is similar to that empirically observed for A/H1N1pdm09 infections.

There are limitations to our study. First, we used clinical observations on viral load dynamics, treatment effectiveness and resistance prevalence measured for seasonal influenza virus infections to parameterize our model. While a future pandemic virus can have distinct epidemiological properties, in contrast to extrapolations considered in previous studies, we sought to inform antiviral treatment demand and impact using a model that is grounded on up-to-date, high-quality empirical data. Second, our deterministic estimates of mean deaths averted represent the central expected impact value for each antiviral. There is considerable variability in observed viral load dynamics and transmission reduction benefit reported in clinical trials such that the true impact of BXM could be lower than our estimates. Nonetheless, our sensitivity analyses consistently demonstrated that BXM could likely reduce more transmissions, and thereby yield greater deaths averted compared to OSL in spite of the uncertainty. Third, although we accounted for imperfect sensitivity of rapid influenza diagnostic tests, we did not consider the impact of false positives on antiviral demand and mortality reduction given uncertainties on background influenza-like illness activity that might prompt uninfected individuals with similar symptoms to seek test-and-treat. However, given the uniformly high specificities of existing rapid tests (>98%) (45), false positives are unlikely going to change our antiviral demand and impact estimates substantially. Finally, our country-level modelling framework and results are intended to inform public health planning and distribution of antiviral stockpiles for individual countries. We have also only considered the impact of drug resistance spread owing to endogenous emergence of resistant mutations within the country and not by exogeneous introduction. Future work should investigate the impact of global resistance transmission dynamics on antiviral effectiveness.

Influenza antivirals, particularly BXM given its demonstrated effectiveness in reducing transmission, have the potential to substantially lower the disease burden of a future influenza pandemic but only if stockpiled in advance and deployable at speed. Our country-specific demand and impact estimates provide a concrete basis for national stockpile planning, regional procurement negotiations, and manufacturer capacity decisions. Yet, no single country can stockpile its way out of a global pandemic emergency. The disproportionate burden falling on lower-income countries, combined with the patent and cost constraints that limit BXM access, means that the life-saving potential our modelling demonstrates will remain unrealised for the populations who need it most, unless the inequities in access to pandemic medicines are resolved before the next pandemic begins.

## Materials and Methods

### Multiscale model

We developed a discrete-time, age-structured, deterministic multi-scale model of influenza transmission that can be flexibly parameterised for individual countries using country-specific demographic and age-dependent contact rate data. Key aspects of our model are detailed below, with the complete formulation provided in the SI Appendix.

To compute transmission dynamics, we adapted the multi-type renewal equation (46) to account for heterogeneity in transmission due to changes in infectiousness distribution by antiviral treatment, age-dependent contact rates between infected and uninfected individuals, susceptibility and protection from infection by PEP of the uninfected individuals. The effectiveness of antivirals predicates on which treatment, when the treatment was initiated since infection. Assuming that the population is stratified into *n* age groups and that *infected* individuals are further distinguished by their diagnosis status and infectiousness distribution *y* as determined by if and when they were administered antivirals, the mean incidence for individuals in group *a* on day *t* is modelled as:

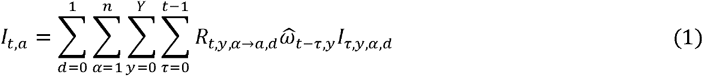

where *R*_*t,y,α* → *a,d*_ is the mean number of secondary infections in age group *a* caused by an infector in age group *α* with infectiousness distribution *y* on day *t* with the boolean status *d* indicating if their infection is identified either by testing or contact tracing; *I*_*t,y,α,d*_ is the number of infected individuals in age group *α* with infectiousness distribution *y* on day *t* with status *d*; and 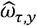 is the normalised infectiousness distribution *y*.

The infectiousness distribution *y* of infected individuals was estimated by: (1) a target cell-limited within-host model (47) that estimates the day-to-day viral load to determine the relative infectiousness of infected individuals over the infectious period, and (2) a correlative model that the links total infectiousness to viral load data given the expected transmission risk reduction conferred by BXM (SI Appendix). Assuming that the pandemic IAV has similar within-host virus replication dynamics to seasonal IAVs, we estimated the cell infection and viral replication rate, as well as treatment effectiveness parameters of antivirals by fitting our within-host model to the viral load measurements taken from clinical trial for different treatment regimens (i.e. placebo, BXM or oseltamivir) that were administered within two days after symptom onset, with (35) and without the emergence of BXM-resistant viruses (6). Given that we still lack a quantitative understanding of how within-host viral load links to between-host transmission,(48) we applied three different correlative models that were previously applied to infer total infectiousness from viral load data of respiratory viral pathogens, estimating the best-fit parameters that align with empirical effectiveness estimates of BXM reducing onward transmission risk by 29% within five days after treatment (7). As all correlative models yield similar transmission dynamics (see SI Appendix for model validation against US influenza season in 2017/2018), we used the Hill equation model for all simulations and applied the best-fit parameters to estimate the mean infectiousness of infected individuals who were treated at different times during the course of their infection (i.e. within and beyond two days after symptom onset).

The timing of treatment administration since infection would depend on their time to symptom onset since infection and delays in treatment since symptom onset. The probability that an infected individual is tested at time *τ* since infection can be formulated as (49):

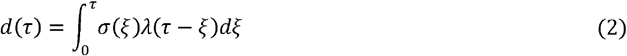

where *σ*(*τ*) is the probability of symptom onset on day *τ* since infection and *λ*(*ν*) is probability of symptomatic testing on day *ν* since symptom onset. 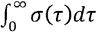 can be less than one and is equal to the probability of a symptomatic infection: 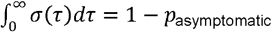 where *p*_asymptomatic_ is expected probability of asymptomatic infection which we assumed to 16% for influenza infections.(50) On the other hand, 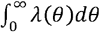 is equal to the willingness of testing provided that all individuals who seek testing will be tested which can also be less than one. In turn, the expected number of infected individuals *T*(*t*) that are positively diagnosed at time *t* is:

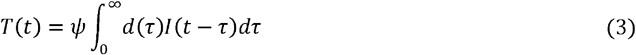

where *ψ* is the sensitivity of the diagnostic test.

In the discrete form, assuming that *d*(*τ*) is the same for all age groups, the expected number of infected individuals of age group *a* with infection age of *τ* days that are positively tested on day *t* is therefore:

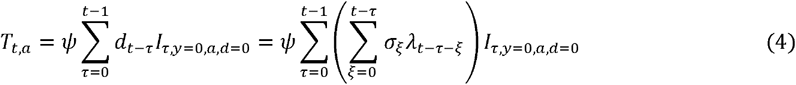

We assumed the ideal scenario where rapid diagnostic tests are used, with no delay in obtaining results after testing, and are widely available and accessible to the whole population (i.e. no shortages).

Infected individuals that test positive may then be given antiviral that will change their infectiousness distribution *y* depending on the infection age *τ* when the antiviral was administered treated and whether or not the individual adhered to the treatment regime. The number of test-and-individuals (*O*_*treatment,τ,a*_) of age group *a* with infection age of *τ* days who are treated is then:

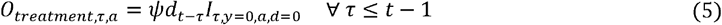

We also assumed that there is no delay in administration of antivirals upon receiving testing results (i.e. testing and drug administration occurs during the same clinical visit).

We assumed that the expected number of deaths correlate with the number of infected individuals. The number of deaths in group *α* on day *t*(*D*_*t,α*_) is

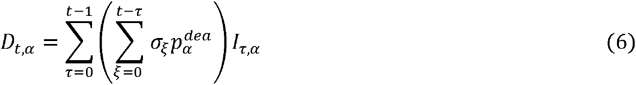

where 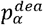 is the probability that symptomatic individuals in group *α* that died.

We included a table of parameters and corresponding values used (Table S1), and a summary of key modelling assumptions (Table S2) made for the between-host transmission model in the SI appendix.

### Antiviral demand

We computed the potential mean antiviral demand for treatment, using either oseltamivir or BXM as the sole infection control intervention, in 186 countries where age demography and contact rates were estimated by the UN World Population Prospects (29) and (51) respectively, during the first wave of four influenza pandemic scenarios with distinct basic reproduction number, age-stratified susceptibility to infection and case fatality rates (i.e. a severe pandemic with a W-shaped age-specific mortality curve, 1918 A/H1N1 (52, 53); a moderately severe pandemic U-shaped age-specific mortality curve, 1968 A/H3N2 (52, 53); a mild pandemic with greater relative mortality rates among adults <65 years of age, 2009 A/H1N1 (30); a severe pandemic with greater relative susceptibility and mortality rates among adults ≥65 years of age, COVID-19 (54– 56); Table S1) akin to the 1918 A/H1N1 (*R*_0_=2.0), 1968 A/H3N2 (*R*_0_=1.8), 2009 A/H1N1 (*R*_0_=1.5) and an influenza pandemic with COVID-19-like reproduction number (*R*_0_=3.0) and disease burden but with IAV-like generation interval. We assumed that 84% of infections were symptomatic even though prevalence of asymptomatic influenza remains largely uncertain (50). We also assumed that asymptomatic individuals are as infectious as their symptomatic counterparts.

To estimate the potential mean antiviral demand, we assumed that there were no age restrictions in access to antivirals and that 95% of all symptomatic individuals would ideally seek clinical intervention within two days after symptom onset, lognormally distributed across time (i.e. mean=1 day, sd=0.6 days after symptom onset). We also assumed that an idealised public health infrastructure was in place such that country-wide distribution of antivirals would begin one week after the pandemic was initiated with ten infections in the country. Given likely limited BXM supply, heterogeneity in healthcare-seeking behaviour, and delays to begin treatment distribution in reality, all of which would lead to lower antiviral demand, we would relax the aforementioned assumptions in subsequent analyses (see below).

Rapid testing would be required for same-day antiviral administration and was performed by a moderately sensitive rapid diagnostic test (70%) (45). Given its multi-day, five-dose treatment course, probability of adherence to oseltamivir treatment was assumed to be lower (65%) (57) than the single-dose BXM (95%). Treated infected individuals that did not adhere to treatment were assumed to be effectively untreated. We estimated the impact of antiviral use by computing the total deaths averted relative to no infection control. Total deaths during the pandemic wave was computed based on net incidence and the corresponding pandemic case fatality rates while accounting for mean reduction in mortality risks of treated individuals by 23% for both oseltamivir (58) and BXM, for which robust mortality risk reduction estimates is currently lacking.

### Additional simulated scenarios

We then used our model to simulate scenarios that deviate from the aforementioned idealised reference case. We simulated different delays to test-and-treatment distribution since pandemic initiation (i.e. one week, four weeks and three months). We also accounted for individual variation in time to treatment since symptom onset (i.e. mean (sd) = 1.0 (0.6), 2.0 (1.2) and 5.0 (3.0) days, which correspond to 95% likelihood of seeking test-and-treat within two, four and eight days since symptom onset respectively). Furthermore, we simulated different BXM distribution strategies, including extending antiviral treatment window from two to eight days, providing PEP to asymptomatic, undiagnosed symptomatic and uninfected household members of positively-tested individuals. While asymptomatic and undiagnosed symptomatic individuals would lower their infectiousness by PEP similarly to symptomatic individuals treated with antivirals, uninfected individuals would lower their likelihood of infection by 57% for ten days after administering BXM as PEP (31). We also simulated scenarios where BXM supply is limited and thus rationed for those aged 5-24 years, 25-64 years or ≥65 years only while the rest of the population were given oseltamivir. Finally, we investigated the impact of BXM resistance emergence and spread based on age-stratified average treatment-emergent resistance prevalence reported for A/H1N1pdm09 (<12 years = 9.4%; ≥12 years = 2.2%) (6, 59, 60) and A/H3N2 (<12 years = 21.0%; ≥12 years = 8.5%) (35, 59, 61) infections. BXM treatment has no effect on within-host replication of BXM-resistant viruses, and in turn their infectiousness. Given limited evidence that BXM-resistant virus may be fitter than their drug-sensitive counterparts (62–64), we conservatively assumed that BXM-resistant strains have similar within-host viral replication fitness and between-host transmission advantage to untreated drug-sensitive viruses (see SI appendix on adjustment of the model to account for relative infectiousness of BXM-resistant variant). We used a Bayesian multilevel model that partially pooled simulation results across all simulated countries and pandemic scenarios to estimate the effect size of each factor on the deaths averted and BXM demand. See SI Appendix for the detailed formulation of the Bayesian multilevel model and assumed priors.

## Acknowledgments

A.X.H., K.D.H. and C.A.R. were supported by the European Research Council NaviFlu (grant 818353). A.X.H. and C.A.R. were also supported by the Dutch Research Council (NWO) Modelling for Pandemic Preparedness grant (10710062310004). A.X.H. was supported by the Dutch Research Council (NWO) Veni Award (09150162210121). C.A.R. was supported by the Dutch Research Council (NWO) Vici Award (09150182010027).

## Author Contributions

A.X.H. and C.A.R. contributed to conceptualization and funding acquisition. A.X.H. contributed to methodology, formal analysis and data curation, and wrote the original draft. A.X.H., K.D.H. and C.A.R. contributed to investigation, validation and visualization. C.A.R. provided supervision. All authors reviewed and edited the manuscript.

## Data and code availability

All simulation data, model source and custom analysis codes are available at https://github.com/AMC-LAEB/flu-antiviral-stockpile.

