## Supplementary material for "The global demand and potential public health impact of oral antiviral treatment stockpile for influenza pandemics": SI Appendix

### Mathematical model

#### Within-host model

We used a within-host model (Figure S8) to estimate the replication dynamics of the inoculating treatment-susceptible virus and treatment-resistant mutant virus since infection, as well as the effectiveness of antiviral treatment to suppress treatment-susceptible virus replication ($e_{treatment}$):

| $\frac{dT}{dt}=-T\left( \beta_{w}V_{w}+\beta_{m}V_{m} \right)\mathcal{-l}TF+rR$ | $(1)$ |
| --- | --- |
| $\frac{dR}{dt}\mathcal{=l}TF-rR$ | $(2)$ |
| $\frac{dI_{\mathcal{V}}}{dt}=\beta_{\mathcal{V}}TV_{\mathcal{V}}-d_{C}I_{\mathcal{V}}-d_{I}I_{\mathcal{V}}F \forall\mathcal{V\in}\left\{ w,v \right\}$ | $(3)$ |
| $\frac{dF}{dt}=p_{F}\left( I_{w}+I_{m} \right)-d_{F}F$ | $(4)$ |
| $\frac{dV_{w}}{dt}=p_{w}\left[ 1-\mu\delta\left( t\geq t_{treatment} \right) \right]\left[ 1-e_{treatment}\delta\left( t\geq t_{treatment} \right) \right]I_{w}-c_{w}V_{w}$ | $(5)$ |
| $\frac{dV_{m}}{dt}=p_{m}\mu\delta\left( t\geq t_{treatment} \right)I_{m}-c_{m}V_{m}$ | $(6)$ |

where $T$, $R$, $I_{w}$, $I_{m}$, $F$, $V_{w}$ and $V_{m}$ are the respective number of uninfected target cells, the number of cells that enter an antiviral state in which they are refractory to infection, the number of cells infected by the wild-type virus, the number of cells infected by the antiviral-resistant mutant virus, the level of interferon response, the amount of free wild-type and mutant viruses. The parameters $\beta_{\mathcal{V}}$, $\mathcal{l}$, $r$, $d_{C}$, $d_{I}$, $p_{F}$, $d_{F}$, $p_{\mathcal{V}}$ and $c_{\mathcal{V}}$ refer to the respective target cell infection rate by variant virus $\mathcal{V}$, interferon-induced antiviral efficacy, reversion rate from antiviral state, virus-induced infected cell death rate, interferon-induced infected cell death rate, production rate of interferons, decay rate of interferons, replication rate and free virus death rate of variant virus $\mathcal{V}$ respectively. $\delta\left( t \right)=1$ when and after the antiviral was administered at time $t_{treatment}$, and is otherwise zero.

We fitted the within-host model to viral load data measured since initiation of different treatment regimens (i.e. placebo, baloxavir marboxil (BXM) or oseltamivir) with and without the emergence of BXM-resistant viruses from a phase 3 clinical trial (CAPSTONE-1) (1, 2). For the BXM-resistance, the rebound in viral load was dominated by BXM-resistant virions based on the estimated within-host mutation frequencies (1). To fit the model, first, we fitted pooled data of the time of symptom onset since influenza virus infection to a lognormal distribution (i.e. mean = 0.35, standard deviation = 0.41; Figure S9A) (3). Second, we fitted the time of treatment initiation since symptom onset reported in the clinical trial to a Gamma distribution (i.e. placebo, shape = 2.91, scale = 8.31; BXM, shape = 2.40, scale = 10.12; oseltamivir, shape = 2.87, scale = 8.53; Figure S9B-D). We then simulated the number of individuals assigned to each treatment regime as per the CAPSTONE-1 trial (i.e. $n=$ 210, 427 and 377 individuals administered with a course of placebo, BXM and oseltamivir respectively (2); $n=$ 34 individuals with treatment-emergent BXM-resistant mutant viruses) (1), randomly drawing the time of symptom onset and treatment initiation from the aforementioned fitted distributions. The CAPSTONE-1 trial reported the viral load distribution of participants in different treatment regimen each day for 7 days since treatment initiation with no further information on when sample collection was performed each day (2). In turn, we randomly generated the timing when viral samples were collected for each individual assuming a uniform distribution within a 12-hour collection time window each day. We then fitted the within-host model to the viral load dynamics simulated individuals, estimating all parameters except for the initial uninfected target ($U_{0}$) and infected ($I_{0}$) cell populations which were assumed to be $4\times{10}^{8}$ and 0 cells respectively (4). We used differential evolution to iteratively search for parameter values that maximise the joint Gaussian likelihood of the viral load data reported in the CAPSTONE-1 trial (2) and in Uehara et al (Figure S8) (1). Applying these maximum-likelihood parameter estimates to our within-host model, we could then simulate the infectious viral load trajectories for different $t_{treatment}$ (Figure S4).

#### Infectiousness estimates from within-host viral load

To correlate within-host viral dynamics to infectiousness (5), we explored three different models that were previously applied to respiratory viral pathogens to estimate infectiousness ($\omega\left( t \right)$) from the simulated viral load ($V\left( t \right)$) at time $t$:

| Hill equation (4, 6) | $\omega\left( t \right)=\frac{1}{1+\left[ \frac{k_{1}}{\log_{10} \left\{ V\left( t \right) \right\}} \right]^{k_{2}}}$  where $k_{1}$ is the viral load value leading to 50% infectiousness and $k_{2}$ is the Hill coefficient. | $(7)$ |
| --- | --- | --- |
| Logit function (7) | $\text{logit}\left[ \omega\left( t \right) \right]=k_{1}\left[ \log_{10} \left\{ V\left( t \right) \right\}-k_{2} \right]+ k_{3} \forall\log_{10} \left\{ V\left( t \right) \right\}>k_{2}$  $\text{logit}\left[ \omega\left( t \right) \right]=k_{3} \forall\log_{10} \left\{ V\left( t \right) \right\}\leq k_{2}$  where $k_{1}$ and $k_{3}$ are the transmission coefficient and baseline log-odds respectively. | $(8)$ |
| Logarithmic function (8) | $\omega\left( t \right)=1-\exp\left\{ \frac{-\log_{10} \left[ V\left( t \right) \right]}{k} \right\}$  where $k$ is the scaling factor. | $(9)$ |

Assuming that treatment with BXM within two days of symptom onset leads to 29% reduction in infectiousness by five days post-treatment as reported in the phase 3 CENTERSTONE trial (9), we applied each infectiousness model to the infectious viral load trajectory of BXM treatment as estimated by our within-host model (Figure S10) and fitted the $k$ parameters by differential evolution. We then applied the best-fit parameters to the within-host and infectiousness models to estimate the infectiousness $\omega$ of infected individuals who were treated at different times during the course of their infection as a result of differences in time to symptom onset since infection, delays in treatment since symptom onset and whether or not resistant mutant viruses emerge after treatment.

We find negligible difference (<10% in total incidence) between all three infectiousness models when combined with the between-host transmission model (see below) to estimate population transmission dynamics (Figure S11).

#### Bayesian posterior sampling of within-host parameters

To characterise uncertainty in the within-host viral load dynamics, we extended the differential-evolution-fitted ODE model to full Bayesian inference using the affine-invariant ensemble sampler implemented in emcee (10). All within-host parameters were sampled jointly. The log-likelihood was defined as a Gaussian over the observed mean and standard deviation of viral load at each sampling timepoint across the placebo, baloxavir, and oseltamivir treatment arms reported in the CAPSTONE-1 trial (2). An informative Beta(50, 2) prior was placed on $e_{treatment}$ for both baloxavir and oseltamivir while and log-uniform priors were assumed for the rest. The sampler was run with 32 walkers for 5,000 steps. Convergence was assessed via integrated autocorrelation time and effective sample sizes were > 200 for all parameters. Five hundred draws were then randomly selected from the posterior and the resulting infectiousness profiles were propagated through the between-host transmission model to obtain the posterior distribution of epidemic outcomes.

#### Between-host transmission model

We developed a discrete-time (i.e. timestep of one day), multi-type renewal process equation model to compute between-hosts transmissions over time to individuals with different susceptibility, contact rates and infectiousness profile. Assuming that the population is stratified into $n$ age groups and that *infected* individuals are further distinguished by their diagnosis status and infectiousness profile $y$, the mean incidence for individuals in group $a$ on day $t$ is modelled as:

| $I_{t,a}=\sum_{d=0}^{1} \sum_{\alpha=1}^{n} \sum_{y=0}^{Y} \sum_{\tau=0}^{t-1} R_{t,y,\alpha\to a,d}\hat{\omega}_{t-\tau, y}I_{\tau,y,\alpha,d}$ | $(10)$ |
| --- | --- |

where $R_{t,y,\alpha\to a,d}$ is mean number of secondary infections in age group $a$ caused by an infector in age group $\alpha$ with infectiousness profile $y$ on day $t$ with the boolean status $d$ indicating if their infection is identified either by testing or contact tracing; $I_{t,y,\alpha,d}$ is the number of infected individuals in age group $\alpha$ with infectiousness profile $y$ on day $t$ with status $d$; and $\hat{\omega}_{\tau, y}$ is the discretised, normalised infectiousness profile and only depends on profile $y$, given by:

| $\hat{\omega}_{\tau, y}=\left\{ \begin{aligned} \int_{\tau-0.5}^{\tau+0.5} \omega_{y}\left( \tau\right)d\tau\forall\tau\geq2 \\ \int_{0}^{1.5} \omega_{y}\left( \tau\right)d\tau\forall\tau=1 \end{aligned} \right.$ | $(11)$ |
| --- | --- |

where $\int_{0}^{\infty} \hat{\omega}_{y}\left( \tau\right)d\tau=1$. Note that $y=0$ denotes the untreated infectiousness profile.

Given the limited evidence that BXM resistant mutations may be fitter or less fit than their wild-type BXM-sensitive counterparts (11–13), we assumed that there is no difference in either within-host viral replication fitness or between-host transmission advantage between the drug-sensitive and resistant virus. In turn, for individuals who are infected by the drug-resistant variant virus, their infectiousness profiles are similar computed by equation 11 for the untreated infectiousness profile. For individuals who are infected by the drug-sensitive virus but later developed treatment-emergent resistance, given that influenza virus transmission bottleneck is likely tight such that either one variant is transmitted upon an infectious contact (14), we adjusted the infectiousness profiles of the drug-sensitive ($\mathcal{V}_{1}$) and resistant ($\mathcal{V}_{2}$) viruses such that:

$$\hat{\omega}_{\tau, y,\mathcal{V}_{1}}^{'}=\hat{\omega}_{\tau, y,\mathcal{V}_{1}}\left( 1-\hat{\omega}_{\tau, y,\mathcal{V}_{2}} \right)$$

$$\hat{\omega}_{\tau, y,\mathcal{V}_{2}}^{'}=\hat{\omega}_{\tau, y,\mathcal{V}_{2}}(1-\hat{\omega}_{\tau, y,\mathcal{V}_{1}})$$

$R_{t,y,\alpha\to a}$ depends on the remaining number of susceptible individuals in age group $a$ on day $t$ ($S_{t,a}=N_{a}-\sum_{t} I_{t,a}$ where $N_{a}$ is the number of individuals of age group $a$ in the simulated population), the number of susceptible individuals in age group $a$ who were administered antivirals as post-exposure prophylaxis $\rho$ days ago ($S_{\rho,a}^{PEP}$) that reduces the likelihood of infection by $\epsilon_{PEP}$ over a time period of $\gamma_{PEP}$ days (i.e. $\rho<\gamma_{PEP}$), the relative risk of infection $M_{\alpha\to a}$ of susceptible individuals in age group $a$ by infected individuals belonging to age group $\alpha$ (assumed to be constant over time) and the impact of infectiousness profile $y$ on mean infectious period:

| $R_{t,y,\alpha\to a}=\frac{S_{t,a}-\epsilon_{PEP}\sum_{t-\gamma_{PEP}}^{t-1} S_{t,a}^{PEP}}{N_{a}}\cdot M_{\alpha\to a}\cdot\left( 1+\epsilon_{y} \right)\cdot R$ | $(12)$ |
| --- | --- |

where $R$ is the basic reproduction number of the simulated pandemic scenario.

Infected individuals with different infectiousness profile $y$ change their mean infectiousness by a factor $\epsilon_{y}$ given by:

| $\epsilon_{y}=\frac{\int_{0}^{\infty} \omega_{y}\left( \tau\right)d\tau}{\int_{0}^{\infty} \omega_{y=\text{untreated}}\left( \tau\right)d\tau}-1$ | $(13)$ |
| --- | --- |

where $\omega_{y}$ is the unnormalised infectiousness of profile $y$ that was computed by either one of the viral load-to-infectiousness models (i.e. equations (7) – (9)).

$\mathbf{M}$ is the normalised relative risk matrix given by:

| $\mathbf{M}=\left[ \begin{matrix} M_{1\to1} & \cdots& M_{\alpha\to1} \\ \vdots& \ddots& \vdots\\ M_{1\to a} & \cdots& M_{\alpha\to a} \end{matrix} \right]=\frac{\mathbf{C}\boldsymbol{\eta}^{T}}{\rho\left( \mathbf{C}\boldsymbol{\eta}^{T} \right)}$ | $(14)$ |
| --- | --- |

where $\mathbf{C}=\left\{ C_{\alpha\to a} \right\}$ is the mean contact rate matrix between individuals in age group $\alpha$ and individuals in age group $a$, $\boldsymbol{\eta}=\left\{ \eta_{a} \right\}$ is the relative susceptibility of individuals in age group $a$, with the most susceptible group having a value of 1 and $\rho\left( \mathbf{C}\boldsymbol{\eta}^{T} \right)$ is the spectral radius of $\mathbf{C}\boldsymbol{\eta}^{T}$. We used country-specific estimates of $\mathbf{C}$ from (15, 16) and estimates of $\boldsymbol{\eta}$ for seasonal influenza (17), 2009 A/H1N1 pandemic (18) and COVID-19 pandemic (19) from various sources. For the 1918 A/H1N1 and 1968 A/H3N2 pandemics, we assumed $\boldsymbol{\eta}=1$ for all age groups.

#### Test and treat

The probability that an infected individual is tested at time $\tau$ since infection can be formulated as (20):

| $d\left( \tau\right)=\int_{0}^{\tau} \sigma\left( \xi\right)\lambda\left( \tau-\xi\right)d\xi$ | $(15)$ |
| --- | --- |

where $\sigma\left( \tau\right)$ is the probability of symptom onset on day $\tau$ since infection and $\lambda\left( \nu\right)$ is probability of symptomatic testing on day $\nu$ since symptom onset. $\int_{0}^{\infty} \sigma\left( \tau\right)d\tau$ can be less than one and is equal to the probability of a symptomatic infection: $\int_{0}^{\infty} \sigma\left( \tau\right)d\tau=1-p_{\text{asymptomatic}}$ where $p_{\text{asymptomatic}}$ is expected probability of asymptomatic infection which we assumed to 16% for influenza infections.(21) On the other hand, $\int_{0}^{\infty} \lambda\left( \theta\right)d\theta$ is equal to the willingness of testing provided that all individuals who seek testing will be tested which can also be less than one. In turn, the expected number of infected individuals $T\left( t \right)$ that are positively diagnosed at time $t$ is:

| $T\left( t \right)=\psi\int_{0}^{\infty} d\left( \tau\right)I\left( t-\tau\right)d\tau$ | $(16)$ |
| --- | --- |

where $\psi$ is the sensitivity of the diagnostic test.

In the discrete form, assuming that $d\left( \tau\right)$ is the same for all age groups, the expected number of infected individuals of age group $a$ with infection age of $\tau$ days that are positively tested on day $t$ is therefore:

| $T_{t,a}=\psi\sum_{\tau=0}^{t-1} d_{t-\tau}I_{\tau,y=0,a,d=0}=\psi\sum_{\tau=0}^{t-1} \left( \sum_{\xi=0}^{t-\tau} \sigma_{\xi}\lambda_{t-\tau-\xi} \right)I_{\tau,y=0,a,d=0}$ | $(17)$ |
| --- | --- |

We assumed the ideal scenario where rapid diagnostic tests are used, with no delay in obtaining results after testing, and are widely available and accessible to the whole population (i.e. no shortages).

Infected individuals that test positive may then be given antiviral that will change their infectiousness profile $y$ depending on the infection age $\tau$ when the antiviral was administered and whether or not the individual adhered to the treatment regime. The number of test-and-treated individuals ($O_{treatment,\tau,a}$) of age group $a$ with infection age of $\tau$ days who are treated is then:

| $O_{treatment,\tau,a}=\psi d_{t-\tau}I_{\tau,y=0,a,d=0} \forall\tau\leq t-1$ | $(18)$ |
| --- | --- |

We also assumed that there is no delay in administration of antivirals upon receiving testing results (i.e. testing and drug administration occurs during the same clinical visit).

#### Post-exposure prophylaxis

Besides symptomatic testing, contacts of positively-tested individuals may be contact traced and be given antivirals as a post-exposure prophylaxis which has been shown to be effective at reducing transmission risk (22, 23).

We focused on forward contact tracing: exposed contacts were identified by contact tracing after an index case was positively diagnosed (24). The number of individuals of age group $a$ that are traced on day $t$ ($c_{t,a}$) depends on the probability that an index case with infection age $\tau$ was tested positive ($d_{\tau})$, the number of infected individuals of age group $\alpha$ with infection age $\tau$ was undiagnosed before day $t$, the expected number of contacts that could be traced and the relative fraction of contacts in age group $a$ to index cases of age group $\alpha$ ($f_{\alpha\to a}$):

| $C_{t,a}=\sum_{\alpha=1}^{n} \sum_{y} \sum_{\tau=0}^{t-1} \psi d_{t-\tau}\cdot I_{\tau,y=0,\alpha,d=0}\cdot\varepsilon f_{\alpha\to a}$ | $(19)$ |
| --- | --- |

Our mathematical framework does not explicitly model the underlying contact networks among individuals. However, assuming that household contacts are the most convenient individuals that can be traced and be given post-exposure prophylaxis, we could approximate the impact of distributing antivirals to exposed household contacts by assuming $\varepsilon$ as the mean number of household contacts based on estimates from the most recent United Nations World Population Prospects for that country (25). $f_{\alpha\to a}$ is inferred from country-specific estimates of mean contact rate matrix in households from (15, 16) The expected number of susceptible individuals of age group $a$ that are traced to be given post-exposure prophylaxis on day $t$ is then computed as:

| $S_{t,a}^{PEP}=C_{t,a}\left( \frac{S_{t,a}-\sum_{t-\gamma}^{t-1} S_{t,a}^{PEP}}{N_{a}} \right)$ | $(20)$ |
| --- | --- |

In turn, the number of previously infected individuals of age group $a$ who are given post-exposure prophylaxis on day $t$ is equals to $C_{t,a}-S_{t,a}^{PEP}$. We assumed that only undiagnosed and untreated infected individuals (i.e. $I_{\tau,y=0,a,d=0}$), including asymptomatic infected individuals, were given post-exposure prophylaxis which would lower their infectiousness that similarly depended on the age of their infection $\tau$ when they were administered the drug. We assumed that post-exposure prophylaxis are distributed multinomially across individuals with infection age $\tau$, employing the Sainte-Laguë apportionment method for integer allocation.

#### Hospitalization and deaths

We assumed that the expected number of hospitalisations and deaths correlate with the number of infected individuals. The number of individuals $H_{t,\alpha}$ in group $\alpha$ that are hospitalised on day $t$ is:

| $H_{t,\alpha}=\sum_{\tau=0}^{t-1} \left( \sum_{\xi=0}^{t-\tau} \sigma_{\xi}p_{\alpha}^{hosp} \right)I_{\tau,\alpha}$ | $(21)$ |
| --- | --- |

where $p_{\alpha}^{hosp}$ is the probability that symptomatic individuals in group $\alpha$ are hospitalised.

Analogously, the number of deaths in group $\alpha$ on day $t$ ($D_{t,\alpha}$) is

| $D_{t,\alpha}=\sum_{\tau=0}^{t-1} \left( \sum_{\xi=0}^{t-\tau} \sigma_{\xi}p_{\alpha}^{dea} \right)I_{\tau,\alpha}$ | $(22)$ |
| --- | --- |

where $p_{\alpha}^{dea}$ is the probability that symptomatic individuals in group $\alpha$ that died.

Table S1 tabulates the parameters used in the between-host transmission model.

#### Bayesian multilevel model

To determine the impact of delays to population distribution and individual treatment, different antiviral distribution strategies including extending the treatment window period since symptom onset, instituting age restrictions on access to treatment and providing prophylactic treatment to exposed contacts, as well as the spread of antiviral resistance, we used a Bayesian multilevel model that partially pooled simulation estimates of percentage of deaths averted by antiviral use and the amount of antivirals demand relative to population size for a range of the aforementioned factors across countries and pandemic scenarios. We assumed a linear correlation between the predicted mean-centered response variable ($Y_{c,p}$) for pandemic $p$ in country $c$ and predictor variables $X_{k}$:

| $\hat{Y}_{c,p}=\sum_{k=1}^{n_{k}} \left( \beta_{k}+b_{k,c}+b_{k,p} \right)X_{k,c,p}+\left( \beta_{0}+b_{0,c}+b_{0,p} \right)$ | $(23)$ |
| --- | --- |

where $\beta_{k},$ $b_{k,c}$ and $b_{k,p}$ are the respective normalised fixed, country- and pandemic-specific effects of predictor variable $k$ while $\beta_{0}$, $b_{0,c}$ and $b_{0,p}$ are the corresponding fixed, country- and pandemic-specific intercepts. Following Gelman et al.,(26) we used weakly informative Student-*t* priors with three degrees of freedom, mean of zero and standard deviation (s.d.) of 2.5 for all effects (i.e. $\beta_{k}, b_{k,c}$ and $b_{k,p}$) while normal priors with mean of zero and s.d. of one were placed for intercepts ($\beta_{0}, b_{0,c}$ and $b_{0,p}$). Half-normal priors with mean of zero and s.d. of one were used for all standard deviations of effects, intercepts and residuals. For correlation between country- and pandemic-level effects, we used the Lewandowski-Kurowicka-Joe correlation prior with shape parameter of one. Posterior sampling was performed using four MCMC chains, with 1000-iteration burn-in and 1000-iteration of saved posterior samples, using a no-u-turn sampler implemented in the ‘brms’ package (27) in R (28). Chain convergence was determined by checking traceplots, ensuring all Rhat values < 1.05 with sufficient effective sample size (>200).

### Supplementary Figures

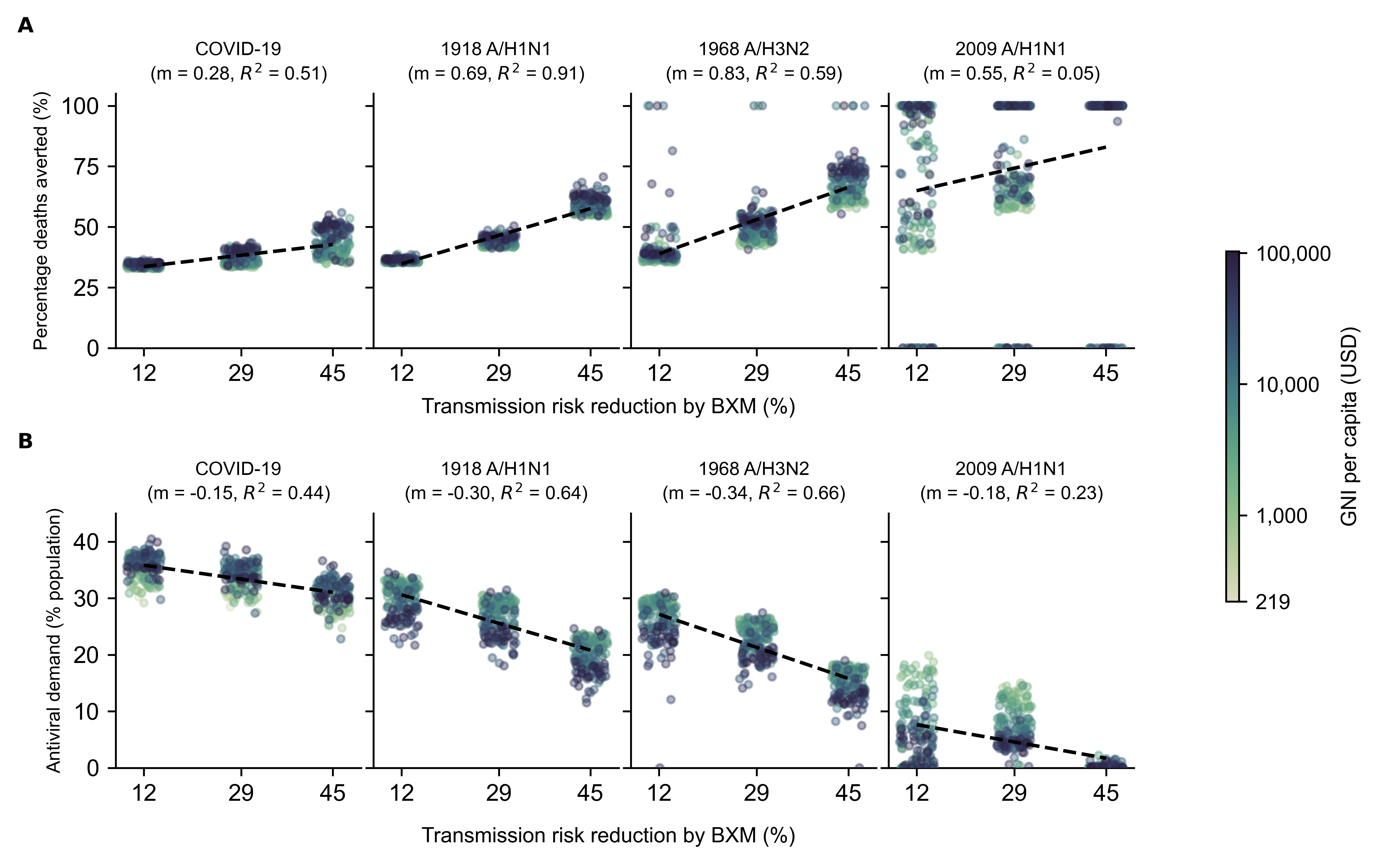

**Figure S1: Sensitivity analysis to account for uncertainty around transmission risk reduction estimate of baloxavir marboxil (BXM).** BXM was administered to all individuals that were positively diagnosed, using a rapid test with 70% test sensitivity, within two days of symptom onset. All symptomatic individuals were assumed to have sought testing within one day after symptom onset on average. Distribution of oral antivirals for treatment began one week after the pandemic was initiated in the country with ten infections. Each circle plot represents a country coloured by its gross national income (GNI) per capita is US dollars (USD) in 2023. Linear regression line is plotted for each pandemic scenario with the estimated slope (m) and coefficient of determination ($R^{2}$) stated in subplot title. (**A**) Mean percentage deaths averted by test-and-treat. (**B**) BXM demand.

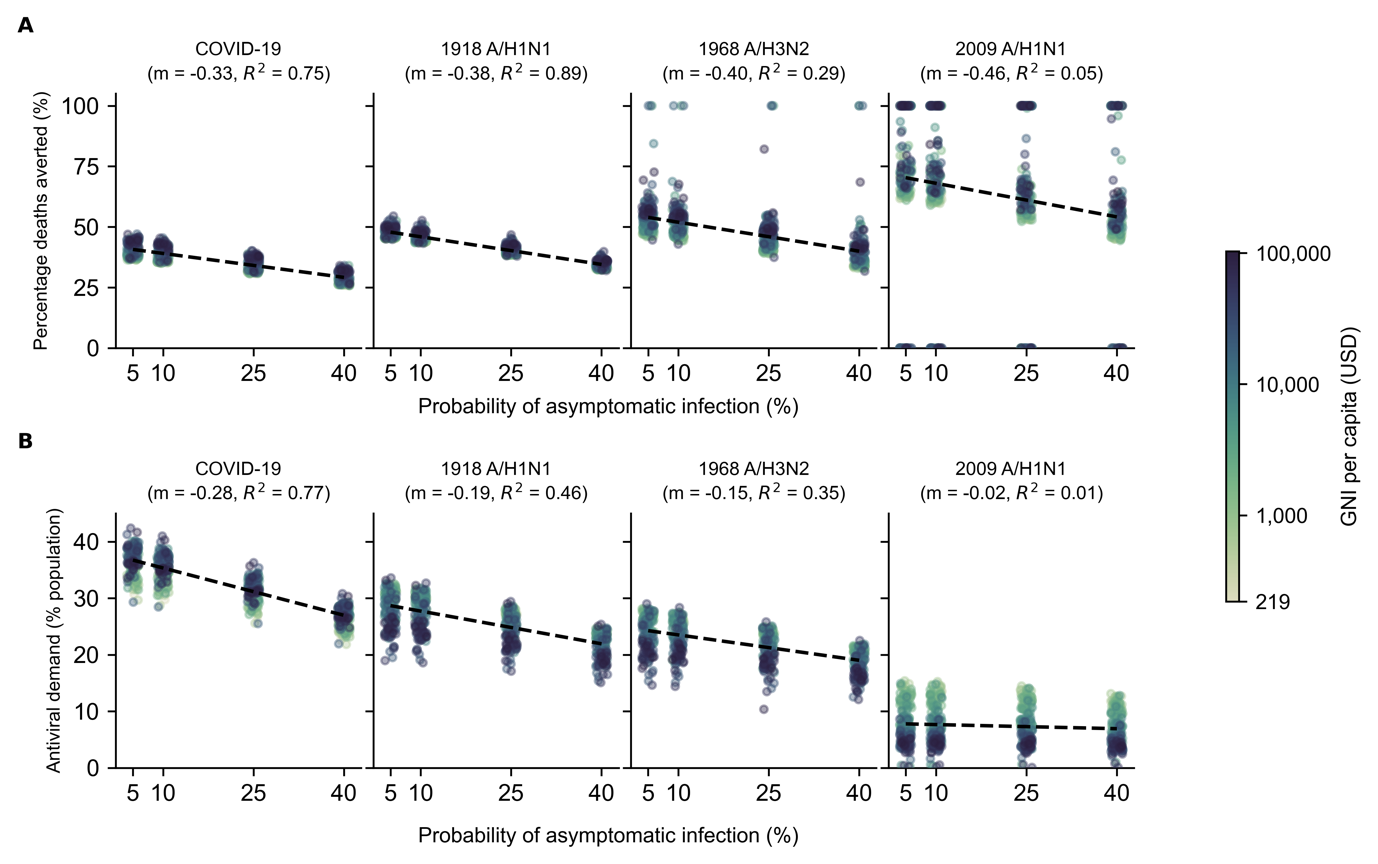

**Figure S2: Sensitivity analysis to account for uncertainty around probability of asymptomatic infection: Treatment with BXM.** BXM was administered to all individuals that were positively diagnosed, using a rapid test with 70% test sensitivity, within two days of symptom onset. All symptomatic individuals were assumed to have sought testing within one day after symptom onset on average. Distribution of oral antivirals for treatment began one week after the pandemic was initiated in the country with ten infections. Each circle plot represents a country coloured by its gross national income (GNI) per capita is US dollars (USD) in 2023. Linear regression line is plotted for each pandemic scenario with the estimated slope (m) and coefficient of determination ($R^{2}$) stated in subplot title. (**A**) Mean percentage deaths averted by test-and-treat. (**B**) BXM demand.

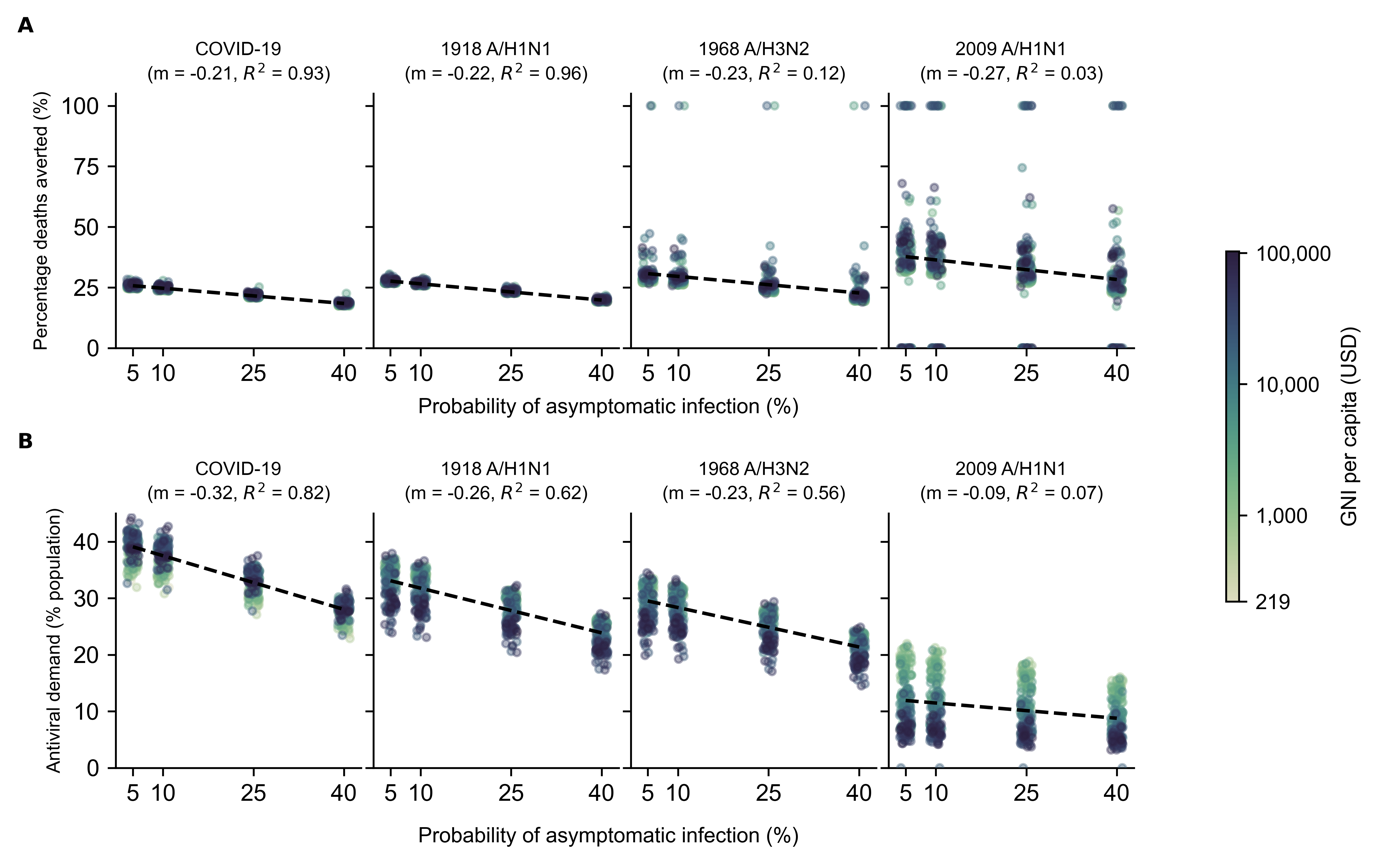

**Figure S3: Sensitivity analysis to account for uncertainty around probability of asymptomatic infection: Treatment with oseltamivir.** Oseltamivir was administered to all individuals that were positively diagnosed, using a rapid test with 70% test sensitivity, within two days of symptom onset. All symptomatic individuals were assumed to have sought testing within one day after symptom onset on average. Distribution of oral antivirals for treatment began one week after the pandemic was initiated in the country with ten infections. Each circle plot represents a country coloured by its gross national income (GNI) per capita is US dollars (USD) in 2023. Linear regression line is plotted for each pandemic scenario with the estimated slope (m) and coefficient of determination ($R^{2}$) stated in subplot title. (**A**) Mean percentage deaths averted by test-and-treat. (**B**) BXM demand.

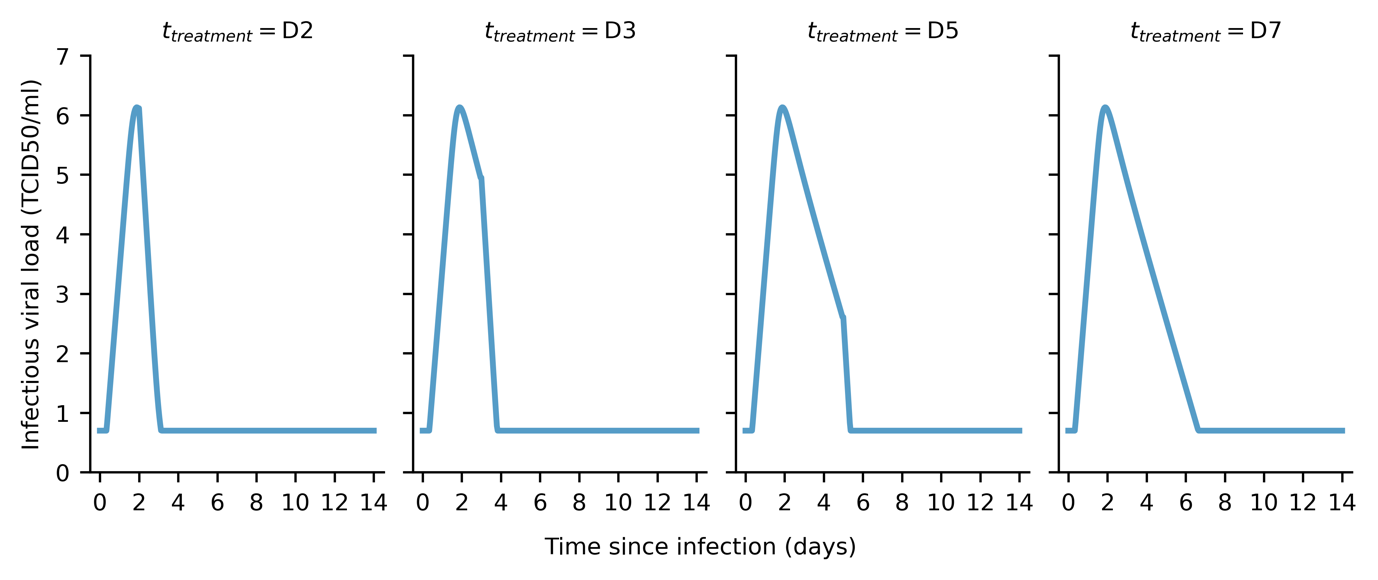

**Figure S4: Estimated infectiousness viral load when treated with baloxavir marboxil at different days since infection (**$\boldsymbol{t}_{\boldsymbol{treatment}}$**).** Within-host viral load trajectories estimated by applying maximum-likelihood parameter estimates to our within-host model and varying $t_{treatment}$.

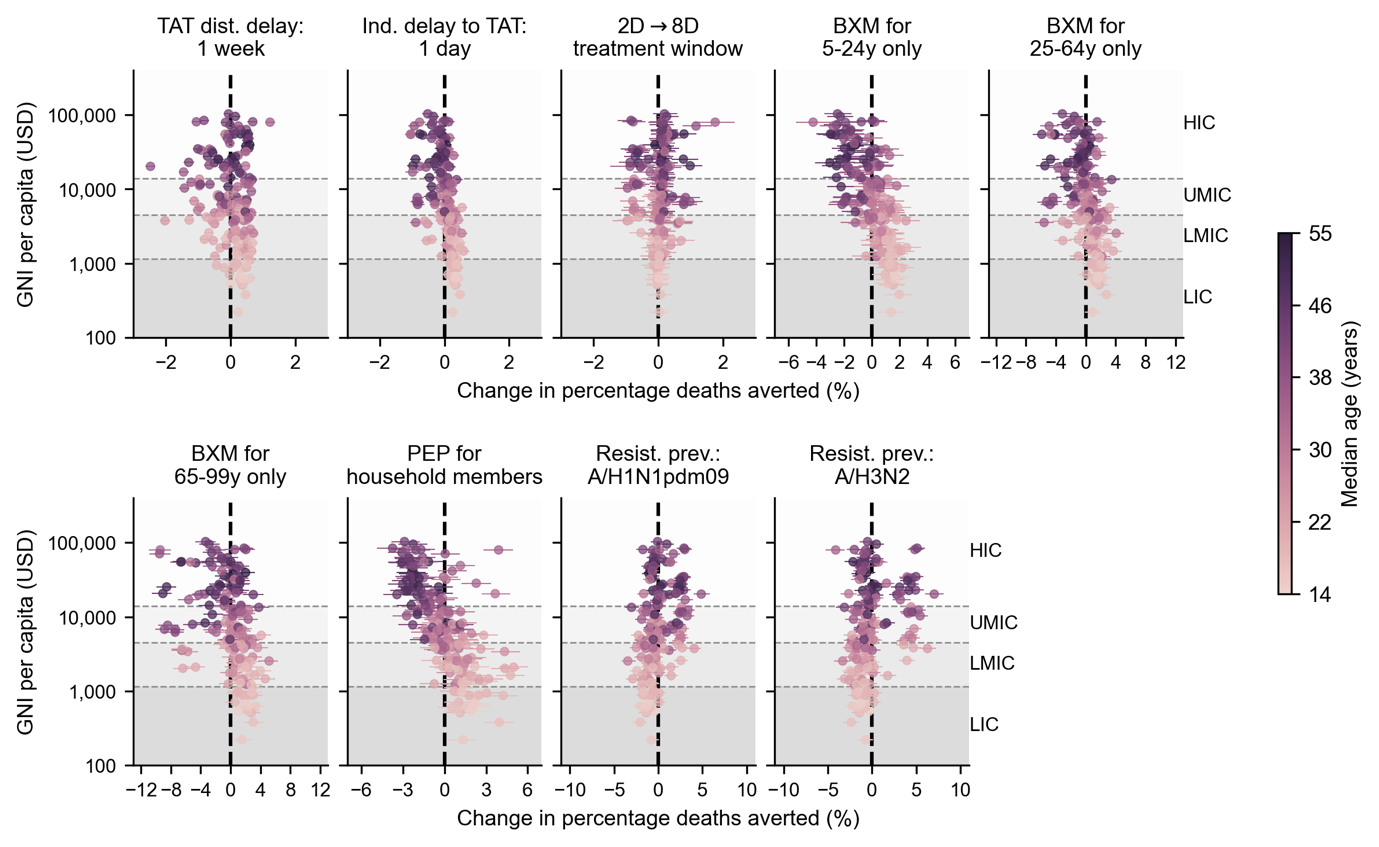

**Figure S5: Country-level effects of treatment delays, distribution strategies, post-exposure prophylaxis and antiviral resistance on mean percentage deaths averted by baloxavir marboxil (BXM) distribution**. Relative to the idealised scenario (i.e. BXM are swiftly distributed to patients across the country one week after the first infection in the country, with no limits on access and availability of tests and antivirals; >95% of all symptomatic individuals are treated within two days after symptom onset if positively diagnosed by a rapid diagnostic test; no emergence and spread of BXM-resistant viruses), mean changes in mean percentage deaths averted due to country-level effects (each circle and line denotes the posterior mean and 95% highest posterior density interval estimated for each country, shading denotes median age of country’s population) are plotted against countries’ gross national income (GNI) per capita is US dollars (USD) in 2023. Background shading denotes the range of GNI per capita distinguishing between different country income groups as classified by the World Bank in 2024 (low income country, LIC; lower-middle income country, LMIC; upper-middle income country, UMIC; and high income country, HIC).

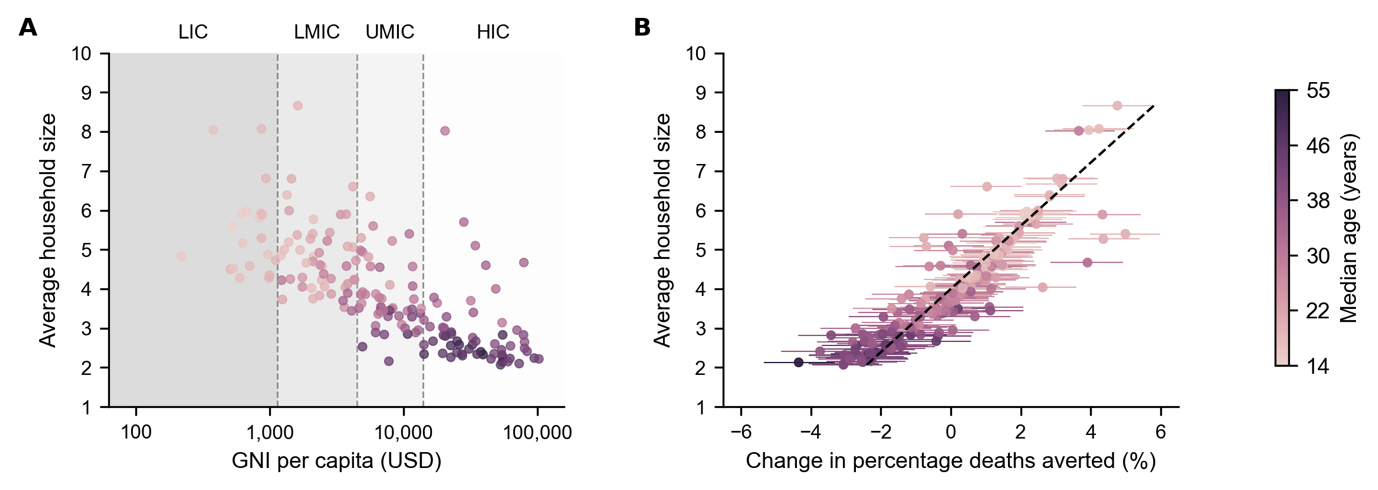

**Figure S6: Correlation between mean household size and country-level effect on mean percentage deaths averted by distributing baloxavir marboxil (BXM) as post-exposure prophylaxis to household members**. (**A**) Mean household size of each country (25) (circle, shading denotes median age of country’s population) plotted against their gross national income (GNI) per capita is US dollars (USD) in 2023. Background shading denotes the range of GNI per capita distinguishing between different country income groups as classified by the World Bank in 2024 (low income country, LIC; lower-middle income country, LMIC; upper-middle income country, UMIC; and high income country, HIC). (**B**) Mean household size of each country (circle, shading denotes median age of country’s population) plotted against mean changes in mean percentage deaths averted due to country-level effects (each circle and line denotes the posterior mean and 95% highest posterior density interval estimated for each country, shading denotes median age of country’s population). Dash line denotes the best-fit linear regression line ($R^{2}=0.80$).

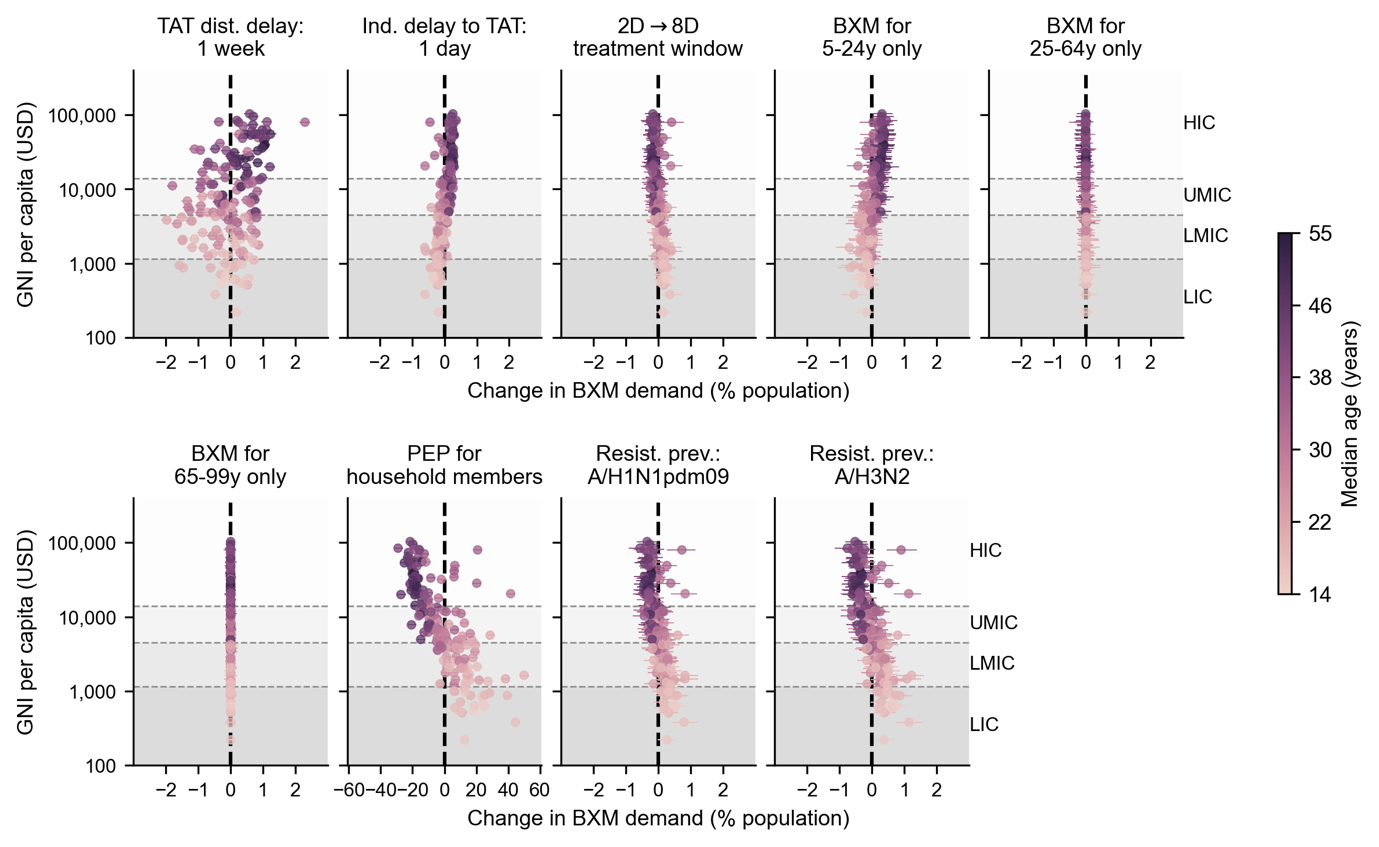

**Figure S7: Country-level effects of treatment delays, distribution strategies, post-exposure prophylaxis and antiviral resistance on mean baloxavir marboxil (BXM) demand**. Relative to the idealised scenario (i.e. BXM are swiftly distributed to patients across the country one week after the first infection in the country, with no limits on access and availability of tests and antivirals; >95% of all symptomatic individuals are treated within two days after symptom onset if positively diagnosed by a rapid diagnostic test; no emergence and spread of BXM-resistant viruses), mean changes in BXM demand due to country-level effects (each circle and line denotes the posterior mean and 95% highest posterior density interval estimated for each country, shading denotes median age of country’s population) are plotted against countries’ gross national income (GNI) per capita is US dollars (USD) in 2023. Background shading denotes the range of GNI per capita distinguishing between different country income groups as classified by the World Bank in 2024 (low income country, LIC; lower-middle income country, LMIC; upper-middle income country, UMIC; and high income country, HIC).

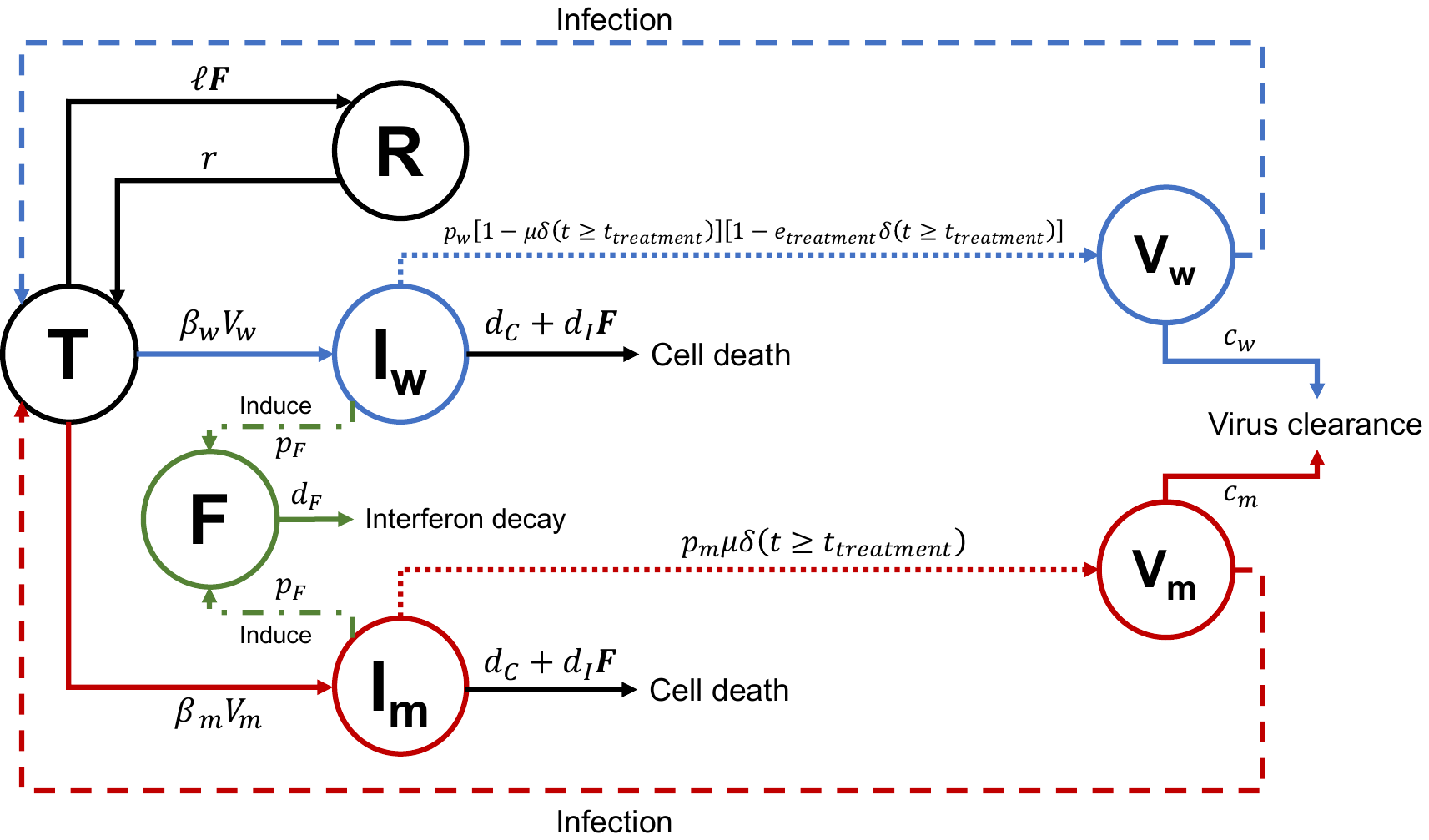

**Figure S8: Within-host model schematic diagram summarizing equations 1-6.**

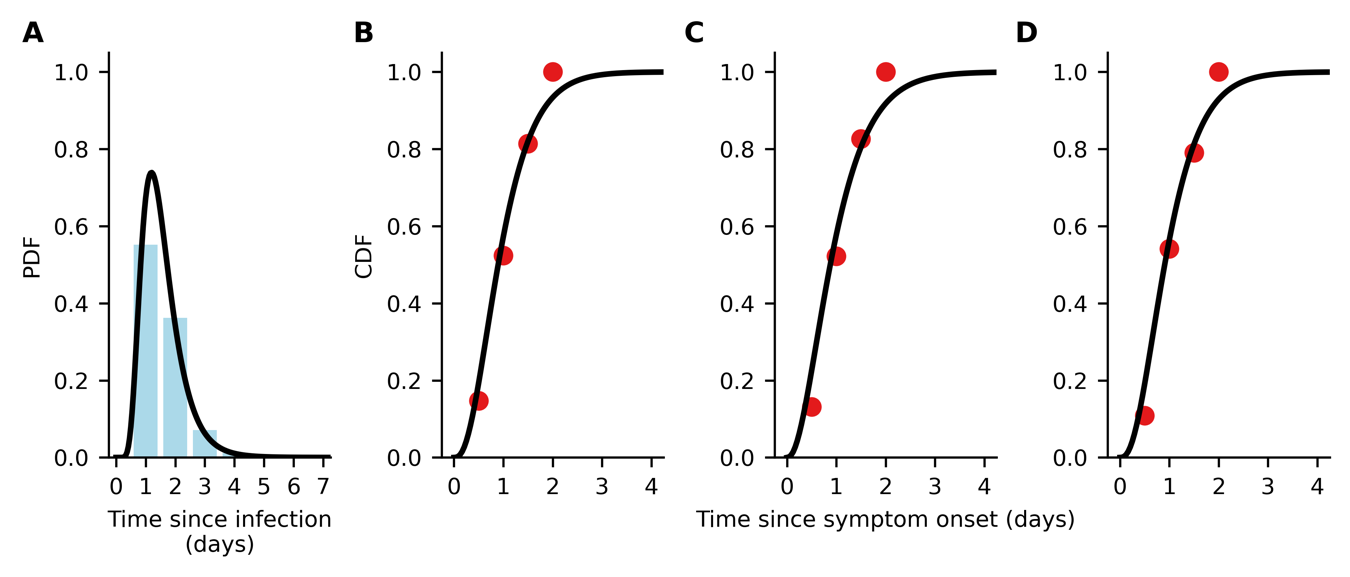

**Figure S9: Data fitting for time to symptom onset since infection and time to treatment administration since symptom onset.** (**A**) Continuous (black line) and discrete (blue bars) probability distribution function (PDF) of lognormal distribution fit to pooled data of the time of symptom onset since influenza virus infection as reported in (3). (**B-D**) Continuous (black line) cumulative distribution function of gamma distribution fit to time of treatment initiation since symptom onset in the phase 3 clinical trial of baloxavir marboxil (red circles) (1, 2); (**B**) placebo, (**C**) baloxavir marboxil, and (**D**) oseltamivir.

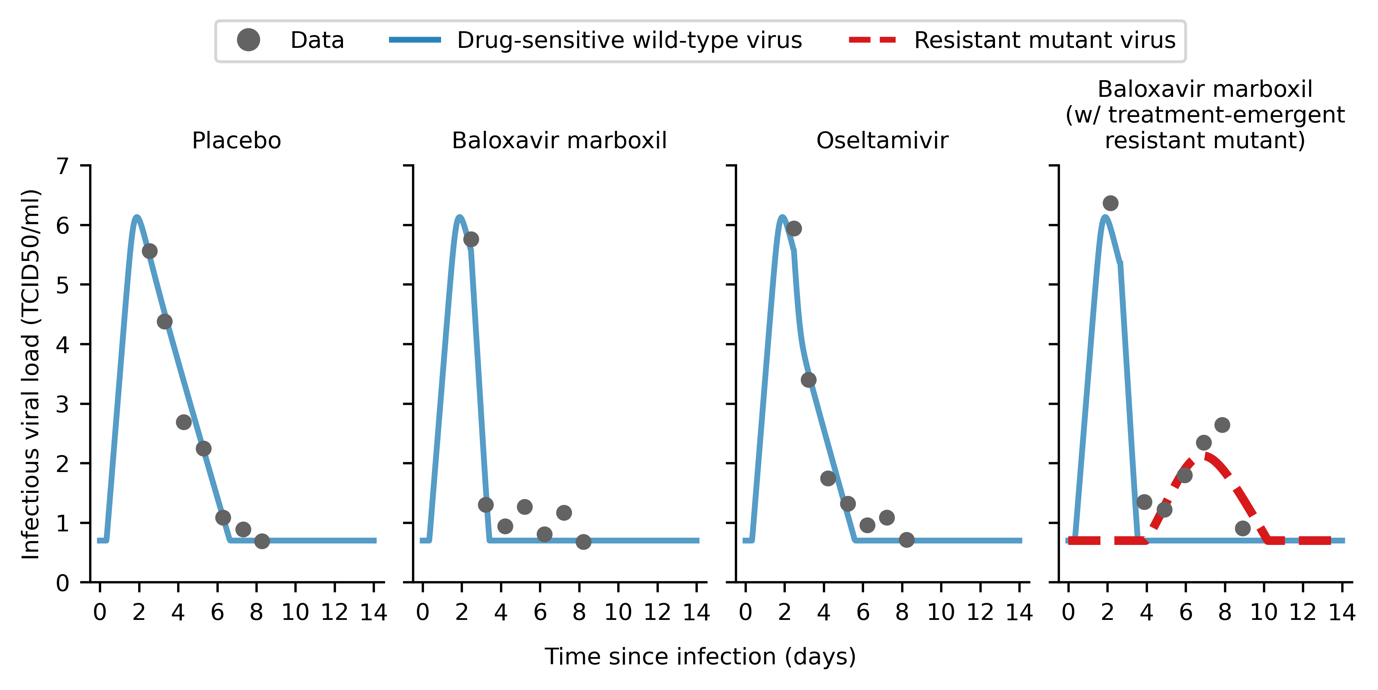

**Figure S10: Within-host viral load model fitted to clinical trial data.** Maximum-likelihood within-host viral load trajectories for drug-sensitive wild type (blue solid line) and drug-resistant mutant (red dashed line) viruses by fitting to mean viral load data (gray circles) obtained in the phase 3 clinical trial of baloxavir marboxil (1, 2).

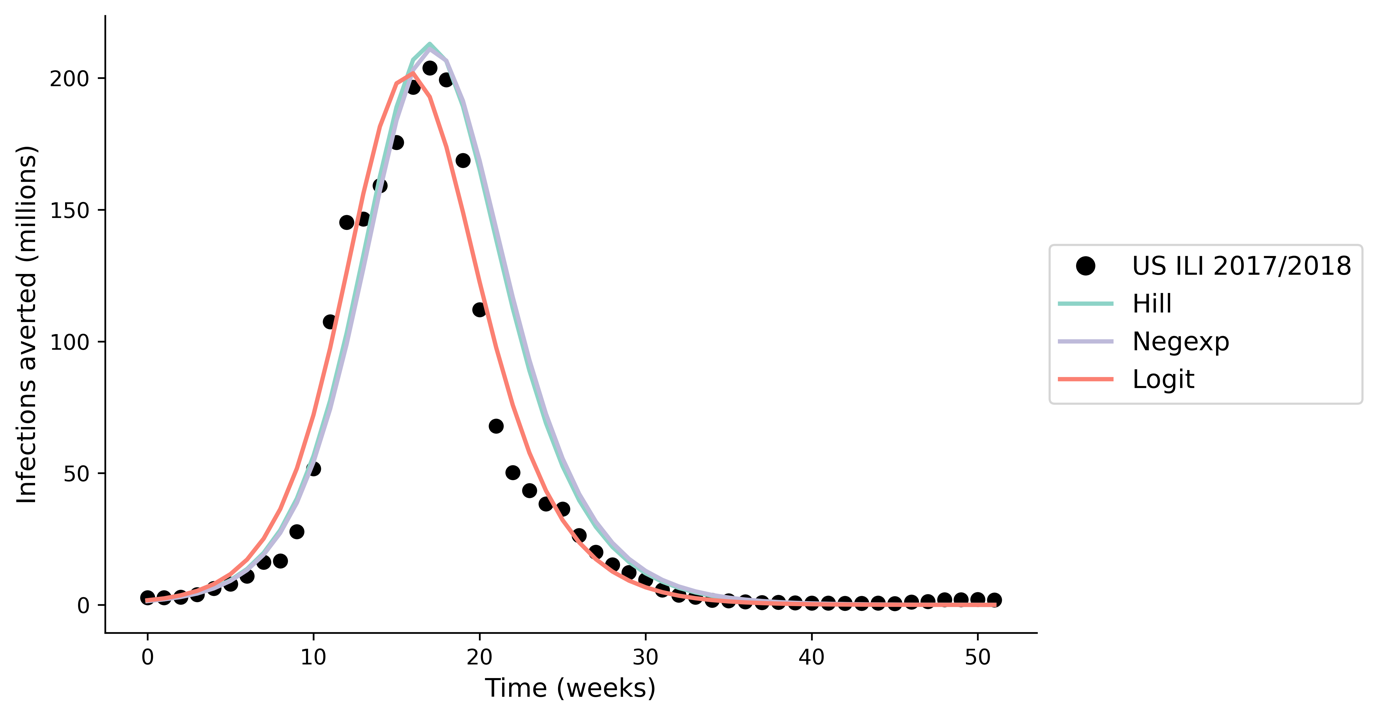

**Figure S11: Model validation; estimated incidence using our multiscale modelling approach when applying different within-host viral load to infectiousness correlative models.** We compared the output of our multiscale model, applying seasonal influenza parameters (17) to estimate the incidence during the 2017/2018 influenza season ($R_{0}=1.3$) (29) in the United States (US), against reported influenza-like (ILI) data (<https://www.cdc.gov/flu/weekly/fluviewinteractive.htm>; black circles). We find minimal difference in total incidence (<10% difference) when applying different within-host viral load to infectiousness correlative models (differently coloured lines).

### Supplementary Tables

**Table S1. Parameters used in the between-host transmission model.**

| **Parameter** | **Value** | **Source** |
| --- | --- | --- |
| Effectiveness in lowering infection likelihood by baloxavir marboxil (BXM) as post-exposure prophylaxis (PEP) ($\epsilon_{PEP}$) | 0.57 | (22) |
| Protective period of BXM as PEP in days ($\gamma_{PEP}$) | 10 |  |
| *Relative susceptibility to infection by age (*$\eta_{a}$*)* | | |
| 1918 A/H1N1, 0-99 years | 1.00 | Assumed |
| 1968 A/H3N2, 0-99 years | 1.00 | Assumed |
| 2009 A/H1N1, 0-19 years | 1.00 | (18) |
| 2009 A/H1N1, 20-64 years | 0.510 |  |
| 2009 A/H1N1, 65-99 years | 0.087 |  |
| COVID-19, 0-14 years | 0.231 | (19) |
| COVID-19, 15-64 years | 0.680 |  |
| COVID-19, 65-99 years | 1.00 |  |
| *Case fatality rate by age (*$p_{\alpha}^{dea}$*)* | | |
| 1918 A/H1N1, 0-4 years | 0.01867 | By fitting W-shape mortality curve reported in (30, 31) |
| 1918 A/H1N1, 5-14 years | 0.00201 |  |
| 1918 A/H1N1, 15-24 years | 0.00719 |  |
| 1918 A/H1N1, 25-34 years | 0.01327 |  |
| 1918 A/H1N1, 35-44 years | 0.00716 |  |
| 1918 A/H1N1, 45-54 years | 0.00512 |  |
| 1918 A/H1N1, 55-64 years | 0.00943 |  |
| 1918 A/H1N1, 65-74 years | 0.03005 |  |
| 1918 A/H1N1, 75-84 years | 0.05380 |  |
| 1918 A/H1N1, 85-99 years | 0.04155 |  |
| 1968 A/H3N2, 0-4 years | 0.0011824 | By fitting U-shape mortality curve reported in (30, 31) |
| 1968 A/H3N2, 5-14 years | 0.0000240 |  |
| 1968 A/H3N2, 15-24 years | 0.0000410 |  |
| 1968 A/H3N2, 25-34 years | 0.0000601 |  |
| 1968 A/H3N2, 35-44 years | 0.0001298 |  |
| 1968 A/H3N2, 45-54 years | 0.0003386 |  |
| 1968 A/H3N2, 55-64 years | 0.0012583 |  |
| 1968 A/H3N2, 65-74 years | 0.0061872 |  |
| 1968 A/H3N2, 75-84 years | 0.0187919 |  |
| 1968 A/H3N2, 85-99 years | 0.0252286 |  |
| 2009 A/H1N1, 0-19 years | 0.00005 | (32) |
| 2009 A/H1N1, 20-64 years | 0.00029 |  |
| 2009 A/H1N1, 65-99 years | 0.00124 |  |
| COVID-19, 0-9 years | 0.001 | (19, 33) |
| COVID-19, 10-19 years | 0.003 |  |
| COVID-19, 20-29 years | 0.012 |  |
| COVID-19, 30-39 years | 0.032 |  |
| COVID-19, 40-49 years | 0.049 |  |
| COVID-19, 50-59 years | 0.102 |  |
| COVID-19, 60-69 years | 0.166 |  |
| COVID-19, 70-79 years | 0.243 |  |
| COVID-19, 80-99 years | 0.273 |  |
| *Basic reproduction number (*$R_{0}$*)* |  |  |
| 1918 A/H1N1 | 2.0 | (34) |
| 1968 A/H3N2 | 1.8 |  |
| 2009 A/H1N1 | 1.5 |  |
| COVID-19 | 3.0 | (35) |
| Probability of asymptomatic infection ($p_{\text{asymptomatic}}$) | 0.16 | (21) |
| Sensitivity of diagnostic test ($\psi$) | 0.70 | (36) |
| Adherence to treatment, BXM | 0.95 | Assumed |
| Adherence to treatment, oseltamivir | 0.65 | (37) |

**Table S2. Key assumptions in the between-host transmission model.**

| No. | Assumption | Implication |
| --- | --- | --- |
| 1 | SIR compartmental model with population structured in 5-year age bins using contact rate matrices estimated by Prem et al. (2021). | Country-level estimation of transmission dynamics with no further spatial delineation. Population contact rates are only structured by age demography. As such, the estimated antiviral demand and impact represent country-level antiviral needs and burden reduction. |
| 2 | Rapid diagnostic testing and antiviral drug administration is assumed to during the same clinical visit. | No delay between obtaining test result and antiviral administration. If there are any delays, the effect would be equivalent to delayed time to seeking test-and-treat, which we had investigated. |
| 3 | No supply and accessibility constraints to diagnostic tests and antivirals. | The key goal of this work is to generate the quantitative evidence to inform antiviral stockpiling efforts prior to the next pandemic. To that end, we focused on estimating the theoretical upper limit of antiviral demand during a nascent influenza pandemic under the ideal assumption of no constraints on diagnostic test and antiviral supply , which in turn informs the capacity needed to meet surge demand. We did, however, investigate the impact of rationing baloxavir marboxil by age and provided estimates on the per-capita amount of diagnostic tests required for each country in Table S3. We also discussed the critical need to expand access to pandemic medicines before the next influenza pandemic in the main text. |
| 4 | High uptake rate (95%) of test-and-treat | Similar to above, we conservatively assumed a high uptake rate of test-and-treat for estimation of the upper limit of test-and-treat demand. We did, however, subsequently investigated how heterogeneity in healthcare-seeking behaviour would lead to heterogenous timing to seeking test-and-treat since symptom onset. |
| 5 | No uptake of pre-pandemic vaccines | We focused on the scenario where the antiviral stockpile was used to limit the impact prior to the availability of vaccines. We have showed in previous work that vaccination is the more resource-efficient intervention compared to test-and-treat during a pandemic (38). |
| 6 | No implementation of non-pharmaceutical interventions (NPIs) | NPIs could lower antiviral demand and augment burden reduction. However, the effectiveness of NPIs are often context dependent, with varying degrees of socio-economic impacts (39) associated with different NPIs that must be carefully considered alongside the cost effectiveness of mass test-and-treat distribution, which is outside the scope of this work. |

**Table S3. Mean deaths averted by test-and-treat with oral antivirals (baloxavir marboxil (BXM) or oseltamivir (OSL), and the corresponding antiviral and rapid testing demand in 186 countries under idealised assumptions.** [Supplementary file]

**Table S4. 95% credible interval of mean percentage deaths averted and mean deaths averted per million people with oral antivirals in the United States.** Credible intervals are generated by first performing Bayesian Markov Chain Monte Carlo sampling to obtain a posterior distribution of all within-host parameters, fitted jointly to the viral load data observed in placebo, BXM and OSL arms reported in Hayden et al (2018). The posterior draws (n=500) are then propagated to the between-host transmission model, simulating for the distribution of BXM and OSL in the United States under the reference pandemic scenarios.

|  | | **Baloxavir marboxil** | | | **Oseltamivir** | |
| --- | --- | --- | --- | --- | --- | --- |
| **Pandemic** | **Mean percentage deaths averted** | | **Mean deaths averted per million people** | **Mean percentage deaths averted** | | **Mean deaths averted per million people** |
| 1918 A/H1N1-like | 19 – 60% | | 1158 – 3,699 | 8 – 40% | | 488 – 2,455 |
| 1968 A/H3N2-like | 10 – 70% | | 60 – 428 | 0 – 45% | | 0 – 279 |
| 2009 A/H1N1-like | 4 – 10% | | 1 – 20 | 0 – 61% | | 0 – 12 |
| COVID-19-like | 24 – 49% | | 1637 – 3,258 | 14 – 33% | | 926 – 2,195 |

**Table S5. Breakdown of baloxavir marboxil (BXM) and oseltamivir stockpile when performing age-based rationing of BXM.** Countries are differentiated by their income groups (low-income, LIC; lower-middle income, LMIC; upper-middle income, UMIC; and high-income, HIC). The median and interquartile range (IQR) of the percentage of stockpile comprising of BXM, and the corresponding per-capita BXM and oseltamivir demand are tabulated.

| **Country income**  **group** | **Ration age group** | **Pandemic** | **Percentage of stockpile comprising of BXM** | | **Per-capita BXM demand** | | **Per-capita oseltamivir demand** | |
| --- | --- | --- | --- | --- | --- | --- | --- | --- |
|  |  |  | **Median** | **IQR** | **Median** | **IQR** | **Median** | **IQR** |
| LIC | 5-24y | 1918 A/H1N1 | 64.1% | 58.4% - 67.7% | 18.3% | 15.6% - 55.2% | 13.7% | 10.3% - 28.5% |
|  |  | 1968 A/H3N2 | 61.6% | 55.9% - 66.0% | 27.7% | 14.5% - 56.9% | 16.0% | 11.3% - 28.2% |
|  |  | 2009 A/H1N1 | 68.8% | 64.5% - 71.7% | 21.5% | 11.1% - 43.2% | 11.2% | 4.4% - 22.7% |
|  |  | COVID-19 | 67.2% | 53.4% - 93.7% | 14.9% | 0.3% - 52.9% | 14.5% | 0% - 25.8% |
|  | 25-64y | 1918 A/H1N1 | 33.8% | 30.5% - 37.8% | 12.8% | 9.7% - 27.2% | 19.8% | 17.1% - 57.4% |
|  |  | 1968 A/H3N2 | 35.4% | 32.6% - 40.2% | 15.0% | 10.6% - 27.2% | 28.7% | 16% - 58.9% |
|  |  | 2009 A/H1N1 | 29.9% | 27.2% - 33.7% | 10.9% | 4.5% - 21.7% | 22.2% | 12.1% - 44% |
|  |  | COVID-19 | 30.9% | 5.8% - 42.8% | 13.5% | 0% - 24.6% | 16.6% | 0.3% - 55.7% |
|  | >64y | 1918 A/H1N1 | 2.5% | 2.0% - 3.0% | 1.0% | 0.7% - 1.8% | 31.6% | 27.3% - 84.6% |
|  |  | 1968 A/H3N2 | 2.5% | 2.0% - 3.1% | 1.1% | 0.8% - 1.8% | 43.0% | 26.7% - 84.8% |
|  |  | 2009 A/H1N1 | 1.0% | 0.7% - 1.8% | 0.5% | 0.1% - 1.2% | 33.9% | 17.1% - 66.1% |
|  |  | COVID-19 | 2.1% | 0.5% - 3.4% | 1.1% | 0% - 1.7% | 29.5% | 0.3% - 80% |
| LMIC | 5-24y | 1918 A/H1N1 | 56.1% | 51.4% - 60.8% | 16.7% | 12.3% - 44.9% | 15.6% | 11.6% - 31.8% |
|  |  | 1968 A/H3N2 | 55.4% | 51.3% - 59.4% | 16.6% | 12.7% - 44.3% | 15.6% | 12.1% - 30.6% |
|  |  | 2009 A/H1N1 | 66.6% | 60.8% - 72.7% | 13.2% | 8.5% - 28.4% | 5.8% | 3.2% - 18.5% |
|  |  | COVID-19 | 59.8% | 48.3% - 81.5% | 14.0% | 0.1% - 45.1% | 17.2% | 0% - 29.8% |
|  | 25-64y | 1918 A/H1N1 | 40.5% | 36.4% - 44.8% | 14.2% | 10.7% - 30% | 18.3% | 13.6% - 47.1% |
|  |  | 1968 A/H3N2 | 41.2% | 38.0% - 44.4% | 14.2% | 11.2% - 29% | 17.9% | 14.1% - 46.4% |
|  |  | 2009 A/H1N1 | 31.9% | 26.4% - 37.3% | 6.9% | 3.4% - 18.5% | 13.4% | 9.3% - 30.7% |
|  |  | COVID-19 | 37.3% | 15.7% - 46.5% | 15.7% | 0% - 28.1% | 16.1% | 0.1% - 48.4% |
|  | >64y | 1918 A/H1N1 | 3.3% | 2.6% - 4.0% | 1.3% | 0.9% - 2.2% | 31.3% | 24.6% - 77.8% |
|  |  | 1968 A/H3N2 | 3.4% | 2.6% - 3.9% | 1.3% | 1% - 2.2% | 29.6% | 26% - 76% |
|  |  | 2009 A/H1N1 | 0.9% | 0.7% - 2.2% | 0.3% | 0.1% - 1.1% | 20.4% | 12.8% - 49.5% |
|  |  | COVID-19 | 3.4% | 2.2% - 4.8% | 1.4% | 0% - 2.4% | 31.6% | 0.2% - 77.1% |
| UMIC | 5-24y | 1918 A/H1N1 | 45.8% | 40.3% - 50.4% | 13.2% | 8.5% - 30% | 16.8% | 11.2% - 31.8% |
|  |  | 1968 A/H3N2 | 46.5% | 42.1% - 51.0% | 13.3% | 9.8% - 30.2% | 16.3% | 13.3% - 30.2% |
|  |  | 2009 A/H1N1 | 64.0% | 59.6% - 74.0% | 8.1% | 5.6% - 15.2% | 4.0% | 2% - 10.9% |
|  |  | COVID-19 | 47.0% | 36.7% - 61.3% | 11.4% | 0.1% - 33.5% | 22.2% | 0% - 36% |
|  | 25-64y | 1918 A/H1N1 | 49.1% | 45.4% - 53.5% | 14.8% | 10.1% - 28.7% | 14.8% | 9.7% - 31.9% |
|  |  | 1968 A/H3N2 | 48.4% | 45.0% - 52.2% | 14.2% | 11.7% - 27.2% | 14.5% | 11% - 31.9% |
|  |  | 2009 A/H1N1 | 34.5% | 25.2% - 38.4% | 4.0% | 2.2% - 11.1% | 8.7% | 6.4% - 16.9% |
|  |  | COVID-19 | 45.9% | 25.3% - 54.7% | 19.5% | 0% - 32.5% | 14.5% | 0.1% - 38.7% |
|  | >64y | 1918 A/H1N1 | 4.8% | 3.9% - 5.7% | 1.6% | 1.1% - 2.7% | 29.2% | 19.5% - 60.8% |
|  |  | 1968 A/H3N2 | 4.5% | 3.7% - 5.2% | 1.5% | 1.2% - 2.5% | 28.1% | 23.3% - 59.5% |
|  |  | 2009 A/H1N1 | 1.2% | 0.6% - 2.2% | 0.2% | 0% - 0.7% | 12.9% | 8.9% - 28.7% |
|  |  | COVID-19 | 6.5% | 4.2% - 8.8% | 2.5% | 0% - 4% | 32.0% | 0.1% - 68.3% |
| HIC | 5-24y | 1918 A/H1N1 | 37.7% | 31.9% - 41.3% | 9.3% | 6.1% - 18.2% | 17.3% | 12.1% - 26.1% |
|  |  | 1968 A/H3N2 | 39.5% | 34.6% - 43.1% | 10.0% | 7.3% - 17.4% | 16.3% | 12.9% - 24% |
|  |  | 2009 A/H1N1 | 66.5% | 61.2% - 77.5% | 5.0% | 3.6% - 6.8% | 2.4% | 1.1% - 4.2% |
|  |  | COVID-19 | 33.8% | 25.6% - 40.7% | 8.5% | 0% - 21.2% | 25.0% | 0.1% - 35.2% |
|  | 25-64y | 1918 A/H1N1 | 55.6% | 52.2% - 59.3% | 14.6% | 10.3% - 22.4% | 10.9% | 7.4% - 20.2% |
|  |  | 1968 A/H3N2 | 53.9% | 51.1% - 57.2% | 13.9% | 11% - 20.5% | 11.2% | 8.4% - 19% |
|  |  | 2009 A/H1N1 | 32.4% | 21.8% - 37.1% | 2.5% | 1.2% - 4.2% | 5.5% | 4% - 7.4% |
|  |  | COVID-19 | 51.1% | 33.3% - 58.8% | 19.9% | 0% - 29.2% | 13.2% | 0.1% - 27.1% |
|  | >64y | 1918 A/H1N1 | 6.6% | 5.3% - 8.2% | 1.8% | 1.2% - 2.7% | 25.2% | 17.1% - 41.5% |
|  |  | 1968 A/H3N2 | 5.9% | 4.7% - 6.7% | 1.5% | 1.1% - 2.4% | 25.2% | 19.8% - 39.5% |
|  |  | 2009 A/H1N1 | 0.5% | 0.3% - 1.5% | 0.0% | 0% - 0.2% | 8.5% | 5.5% - 11.9% |
|  |  | COVID-19 | 12.3% | 8.3% - 17.4% | 3.7% | 0% - 5.7% | 29.3% | 0.1% - 49.5% |
